# An aging focused unobtrusive and Privacy-Preserving Digital Behaviorome

**DOI:** 10.1101/2021.12.23.21267929

**Authors:** Narayan Schütz, Samuel E.J. Knobel, Angela Botros, Michael Single, Bruno Pais, Valérie Santschi, Daniel Gatica-Perez, Philipp Buluschek, Prabitha Urwyler, Stephan M. Gerber, Réne M. Müri, Urs Mosimann, Hugo Saner, Tobias Nef

## Abstract

Digital measures are increasingly used as objective health measures in remote-monitoring settings. In addition to their use in purely clinical research, such as in clinical trials, one promising application area for sensor-derived digital measures is in technology-assisted ageing and ageing-related research. In this context, digital measures may be used to measure the risk of certain adverse events such as falls, and also to provide novel research insights into ageing and ageing-related conditions, like cognitive impairment. While major emphasis has been placed on deriving one or more digital measures from wearable devices, a more holistic approach inspired by systems biology that leverages large, non-exhaustive sets of digital measures may prove highly beneficial. Such an approach would be useful if combined with modern big data approaches like machine learning. As such, extensive sets of digital measures, which may be referred to as digital behavioromes, could help characterise new phenotypes in deep phenotyping efforts. These measures could also assist in the discovery of novel digital biomarkers or in the creation of digital clinical outcome assessments. While clinical research into digital measures focuses primarily on measures derived from wearable devices, proven technology used for long-term remote monitoring of older adults is generally contactless, unobtrusive, and privacy-preserving. In this context, we introduce and describe a digital behaviorome: a large, non-exhaustive set of digital measures based entirely on contactless, unobtrusive, and privacy-preserving sensor technologies. We also demonstrate how such a behaviorome can be used to build digital clinical outcome assessments that are relevant to ageing and derived from machine learning. These outcomes included fall risk, frailty, mild cognitive impairment, and late-life depression. With the exception of late-life depression, all digital outcome assessments demonstrated a promising ability (ROC AUC *≥* 0.7) to discriminate between positive and negative health outcomes, often in the range of comparable work with wearable devices. Finally, we highlight the possibility of using these digital behaviorome-based outcome assessments to discover novel potential digital biomarkers for each outcome. Here, we found reasonable contributors but also some potentially interesting new candidates regarding fall risk and mild cognitive impairment.

## 1 Introduction

Modern sensor technologies are increasingly used to obtain health information in remote-monitoring settings [1]. While no commonly accepted nomenclature exists when referring to sensor-derived health information, we try to adhere to recommendations set out by Goldsack et al. [2, 3]. As such, we use the term ‘digital measure’ to broadly describe any sensor-derived measurement of health. Applications of digital measures in general can be diverse, ranging from providing novel endpoints in clinical trials [4] over healthcare monitoring [5] to wellness monitoring with fitness trackers. Following Goldsack et al., meanwhile, we use the term ‘digital biomarkers’ to refer to individual digital measures that apply an easy-to-grasp concept and are strongly correlated to a known measurement of disease [2, 3]. An example of this is the number of steps per day, as measured by a digital device, which has been shown to be strongly associated with cardiovascular disease and mortality [6]. We use the term ‘digital clinical outcome assessment’ (COA) for digital measures that describe how a patient feels, functions, or survives [2, 3]. This latter type of digital measure may utilise different sensor modalities and other digital measures without necessitating the measurement of an underlying physical concept. For instance, digital measures of gait and finger-tapping might be used to build a machine learning-derived digital Parkinson’s disease severity measure [7].

While much recent interest in this area has centred on the use of digital measures in clinical trials, another traditional field of application has been in technology-assisted ageing and ageing-related research. With the rapid ageing of many countries’ populations, age-related social and economic challenges are starting to gain prominence [8, 9]. In this area, modern information technology may help solve, or at least alleviate, some of the associated negative effects of societal ageing, such as steadily increasing healthcare costs [10, 11]. Moreover, digital measures could provide information about an individual’s functional status and health changes, thereby facilitating earlier and more personalised interventions that may eventually allow older adults to remain independent for longer and increase their quality of life [5]. Although limited to the efforts of just a few research groups, research in this direction has been ongoing for more than a decade, amounting to numerous successful studies around the world [12, 5, 13, 14, 15, 16, 17, 18, 19, 20]. For instance, Rantz et al. show how sensor technologies linked to early alert systems led to better health outcomes among older adults [5]. Furthermore, Hayes et al. demonstrate that variations in sensor-derived gait speed and physical activity differ significantly between older adults with mild cognitive impairment (MCI) and healthy older adults [18]. Meanwhile, in our own research, we recently found that nightly movements in bed may be a promising digital measure; with further validation, these movements could potentially constitute a digital biomarker for monitoring signs of early health deteriorations in older adults [16]. In another line of research, Piau et al. have demonstrated how in-home walking speed may be a potential digital biomarker for fall risk among older adults [21]. Furthermore, Austin et al. report that digital measures were used successfully to assess loneliness in a study with 16 older adults [22]. In addition to directly improving health outcomes by means of early detection of health issues, digital measures in ageing research may also enable novel insights into ageing and ageing-related conditions [23].

Sensor technology used in populations of older adults is commonly unobtrusive or even contactless, with minimal to no participant interaction and the goal to minimises privacy intrusion. These sensor technologies contrast with the predominant use of wearable devices employed to derive digital measures in clinical research. There are multiple reasons for this: (1) older adults tend to be more wary of novel technologies [24]; (2) since monitoring durations are potentially very long or even unlimited, it is ideal if there is no interaction with the system, as there is evidence of wear-time-dependent compliance issues [25]; (3) there is a certain stigma attached to the use of wearable devices, whereby many older adults tend to fear being seen as frail if they wear a device – even if it is just an alarm clock [24]; (4) for users with potential memory issues, wearing and maintaining devices may not be feasible. Due to these factors, most long-term research using sensor technology with older adults has focused on unobtrusive and cost-effective contactless devices [18, 13, 5, 19, 23, 26, 14]. Sensors successfully used in ageing-related, real-world, long-term monitoring projects include passive infrared (PIR) motion sensors that capture an individual’s activity in a given room [5, 18, 13, 19, 23, 27], contact door sensors that can signal when a person leaves or enters the home [15, 26, 23], pressure sensors on or under a mattress that capture sleep measures [16, 23, 5], and electronic pillboxes to track medication adherence [28], along with potentially more obtrusive depth-sensing cameras that track silhouettes to detect falls and gait parameters [5]. A primary downside of using this kind of contactless, unobtrusive technologies, as opposed to wearables, is the additional environmental influences such sensor placement, apartment layouts or material properties. This may lead to more noise measurements as well as potentially worse inter-subject reliability.

Currently, digital measures for ageing and other areas are commonly evaluated individually or by combining several specific measures based on concepts of interest. While this approach is entirely reasonable, it may limit the potential of digital measures. We therefore hypothesise that a more holistic approach — inspired by systems biology and applying digital measures — may be highly promising. This involves using larger sets of digital measures, potentially in the hundreds to thousands, which may be particularly helpful in exploratory research and may enable the creation of strong digital COAs by leveraging large-scale machine learning approaches. This approach is analogous to more classical biological settings, where measurements can assess individual blood tests or genes (for instance, by means of single-nucleotide polymorphisms) as well as whole sets, such as metabolomes or genomes, to identify new phenotypes of health and disease. Since science is primarily informed by what it can measure [29], the introduction of a new layer of information such as this could potentially lead to significant new research insights [30]. In the context of unobtrusive and contactless technologies, this could also help to counteract some of the downsides of those technologies by basing algorithms on a larger set of measures, that may bring about some redundancies.

In the context of digital measures, an extensive set may be called a digital behaviorome, in which the basic measurement unit is a digital measure – as opposed to, for instance, a gene or metabolite in a genome or metabolome, respectively. In this train of thought, Rashidisabet et al. define a digital behaviorome as ‘a subset of a person’s behavior measurable by connected devices via a systems biology approach’ [31]. However, it remains to be seen whether this definition is overly broad, as it would encompass measures for parameters such as vital signs, which are only indirectly related to behaviour. Furthermore, it should be taken into account that, unlike with biological ‘omes’, a complete set is likely never attainable, as there may be an infinite number of possible digital measures to be extracted, such that a digital behaviorome is a relatively non-exhaustive set of digital measures. Two notable real-world examples of using extensive sets of digital measures are studies by Cook et al. [27] and Chen et al. [32], which demonstrated the feasibility of using a behaviorome based on wearable and contactless sensors to predict multiple clinical scores (in the former) and mild cognitive impairment (in the latter).

Building on their work, we introduce a detailed, unobtrusive digital behaviorome, constituting a non-exhaustive set of 1268 existing and novel digital measures. We emphasise the importance of basing these measures on sensor technology that is evidently accepted by older adults, in addition to being affordable and feasible in real-world deployments. Thus, all of these measures are based on a small set of unobtrusive, contactless, and cost-effective sensors that, as shown by Baettie et al., scale well to large ageing-related remote-monitoring projects [23]; thus, these sensors should be compatible with most long-term monitoring projects in community-dwelling older adults.

In this study, we make three major contributions: (1) We introduce and describe the digital measures that comprise the proposed unobtrusive behaviorome (including detailed descriptions as part of accessible supplementary online material); (2) We showcase the potential for using this behaviorome to create ageing-relevant COAs for fall risk, frailty, late-life depression, and MCI; (3) We highlight the possibility of discovering novel digital biomarkers on the basis of the introduced COAs.

## 2 Results

### 2.1 An Unobtrusive Digital Behaviorome

We introduce a set of 94 hypothesis-driven base measures, from which we further derive a total of 1268 digital measures using aggregation and frequency analysis. All measures are obtainable through unobtrusive and privacy-preserving contactless sensors that do not require any participant interaction. Of these 1268 digital measures, 223 were extracted by means of PIR sensors in essential rooms (the entrance, bathroom, living room, bedroom, and kitchen) and magnetic door sensors on the refrigerator and entrance door. An additional 1046 measures were extracted on the basis of sleep data from a quasi-piezoelectric bed sensor placed under the mattress. Detailed descriptions and derivations, as well as associated hypotheses, are provided in the supplementary material, together with a high-level summary table^1^ of all presented digital measures. Furthermore, an extensive online version with interactive visualisations, along with additional data including measure distributions and correlations with various ageing-relevant health indicators and outcomes, is available online at GitHub ^2^ and serves as an online supplementary to this article. An example of averaged behavioromes is shown in Figure 1.

**Figure 1:**
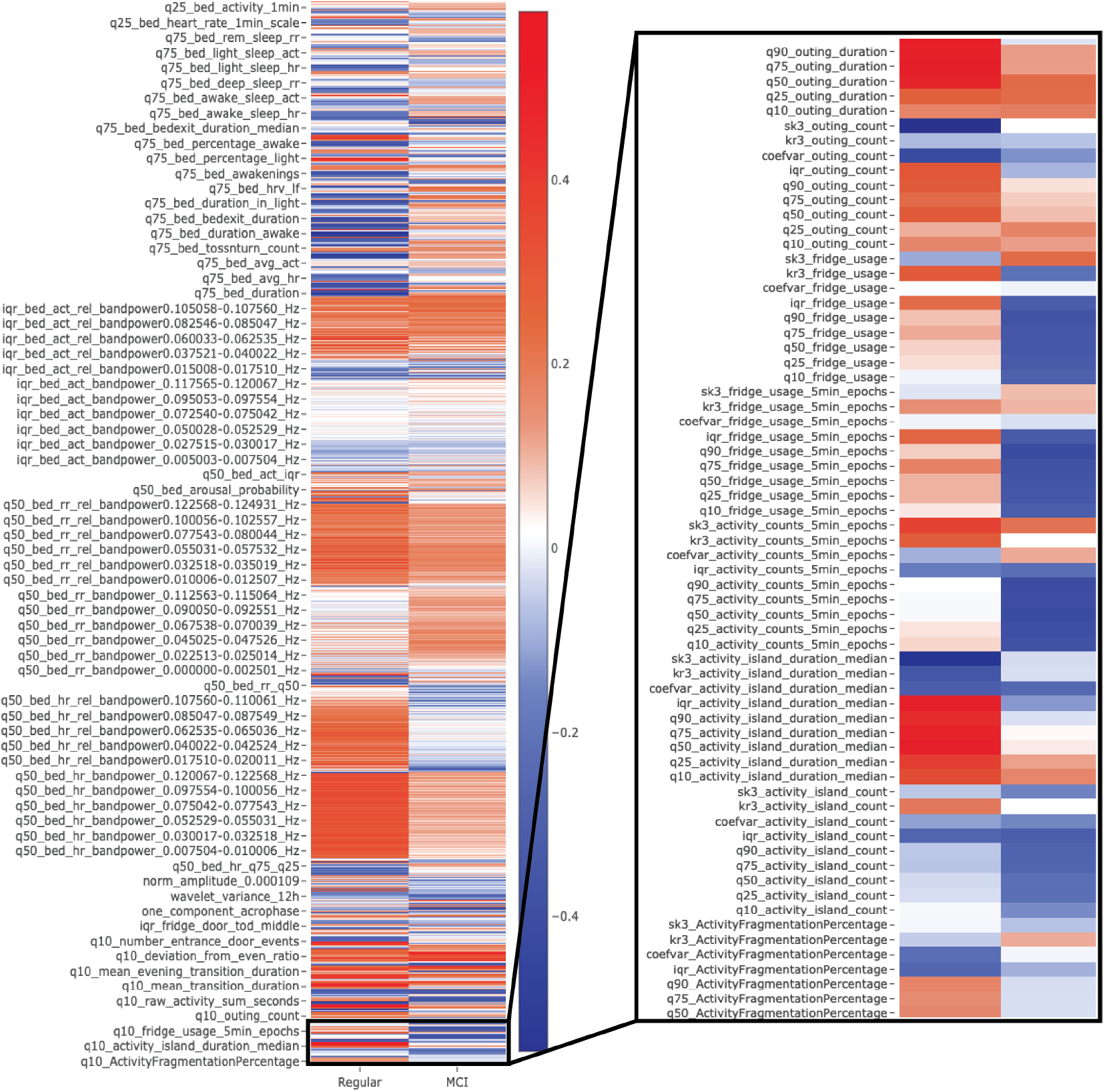
Depicts an example of z-normalised, averaged behavioromes of participants with mild cognitive impairment (MCI) (based on a Montreal Cognitive Assessment screening < 23 points). Digital measures *>* 0 (in blue) indicate above-average values for that group, while *<* 0 (red) indicates below-average values. Many digital measures visually differ in both examples. It should be noted that this is a down-scaled visualisation, as not all measures would fit in the figure. For the complete and interactive version, see the *supplementary online version*.

### 2.2 Machine Learning based Digital Clinical Outcome Assessments

Here, we demonstrate how the introduced digital behaviorome could be useful for for ageing and ageing related research. To this end, we created machine learning-derived digital clinical outcome assessments, aimed at automatically classifying ageing-relevant health outcomes based on the behaviorome. We created five datasets, one based on each clinical assessment, including fall risk, frailty, late-life depression, and MCI. This analysis is based on remote-monitoring data from two observational longitudinal pilot studies in Switzerland, where independently living, community-dwelling older adults were equipped with pervasive computing systems and monitored over the course of a year, while simultaneously being subject to regular visits and clinical assessments. The results on predicting ageing-relevant positive and negative health outcomes are summarised in Table 1. The differences between using the behaviorome alone versus using the behaviorome in addition to demographics were minimal and, judging by overlapping 95% CIs, non-significant. The highest discriminative power, in terms of ROC AUC (area under the receiver-operating characteristic curve), was achieved with the Tinetti Performance-Oriented Mobility Assessment (POMA)-based fall-risk dataset when including both demographic and behaviorome information (ROC AUC = 0.805). Notably, however, demographic information alone was sufficient to produce good performance (ROC AUC = 0.777) in this particular case. Performances on the fall-risk related Timed Up and Go Test (TUG), the MCI-related Montreal Cognitive Assessment (MoCA), and the frailty-related Edmonton Frail Scale (EFS) datasets were also relatively high, with ROC AUC values of 0.786, 0.780, and 0.704, respectively, when using only the behaviorome. The worst performance was achieved with the dataset based on the Geriatric Depression Scale (GDS) (ROC AUC = 0.620), when using only the behaviorome. Here, the difference between using only demographics versus using the behaviorome was also minimal, with a slight but non-significant advantage in favour of the behaviorome-only scenario. Overall, though, the addition of the behaviorome resulted, in all cases, in higher ROC AUC and PrAUC (Area Under the Precision-Recall Curve) values than those obtained when using only demographic information. These differences were statistically significant in all but the POMA and GDS datasets (which were based on rather conservative, non-overlapping CI intervals). The largest differences were found with the MoCA and TUG datasets, for which the objectives were to identify participants with an indication of mild cognitive impairment or increased fall risk, respectively.

**Table 1:** Results for the creation of machine learning-derived digital COAs based on clinical assessments for fall risk (TUG & POMA), frailty (EFS), late-life depression (GDS), and mild cognitive impairment (MoCA). TUG: Timed Up and Go Test POMA: Performance-Oriented Mobility Assessment EFS: Edmonton Frail Scale MoCA: Montreal Cognitive Assessment

| Dataset | ROC AUC (± 95 CI) | | | PrAUC (± 95 CI) | | |
| --- | --- | --- | --- | --- | --- | --- |
|  | Demographics | Behaviorome | Behaviorome + Demographics | Demographics | Behaviorome | Behaviorome + Demographics |
| TUG | 0.662 ± 0.041 | 0.786 ± 0.037 | 0.783 ± 0.036 | 0.681 ± 0.035 | 0.816 ± 0.033 | 0.805 ± 0.034 |
| POMA | 0.777 ± 0.028 | 0.782 ± 0.035 | 0.805 ± 0.027 | 0.615 ± 0.046 | 0.650 ± 0.047 | 0.678 ± 0.044 |
| EFS | 0.564 ± 0.034 | 0.704 ± 0.039 | 0.698 ± 0.038 | 0.523 ± 0.041 | 0.625 ± 0.045 | 0.603 ± 0.044 |
| GDS | 0.602 ± 0.046 | 0.620 ± 0.048 | 0.616 ± 0.048 | 0.397 ± 0.051 | 0.477 ± 0.054 | 0.458 ± 0.053 |
| MoCA | 0.430 ± 0.038 | 0.780 ± 0.039 | 0.757 ± 0.038 | 0.653 ± 0.027 | 0.863 ± 0.028 | 0.839 ± 0.028 |

### 2.3 Evaluation of Individual Digital Measure Importance

The importance of individual digital measures with respect to the COAs were evaluated by means of SHapley Additive exPlanations (SHAP) values [33, 34]. In Figure 2, we present the most important digital measure, based on global SHAP values, across all 100 simulations of each single dataset – corresponding to the different COAs. A more detailed table, highlighting the top 10 highest-ranked measures based on global SHAP values, is available in the Appendix C.III. Furthermore, in Figure 3, we display *beeswarm* plots of SHAP values for the individual COAs. These show how the nine most important digital measures – as well as the sum of all other measures combined – influence the log odds ratio of having a negative health outcome on the various datasets. Values shown in Figure 3 mostly align with the global SHAP importance rankings, although there are minor differences. Since the global SHAP rankings are based on 100 iterations (and thus 100 models), and since the importances shown in Figure 3 are based on a single model, we generally place greater emphasis on the global SHAP values. Nonetheless, the *beeswarm* plots are still useful, as they provide insights into the direction of effects.

**Figure 2:**
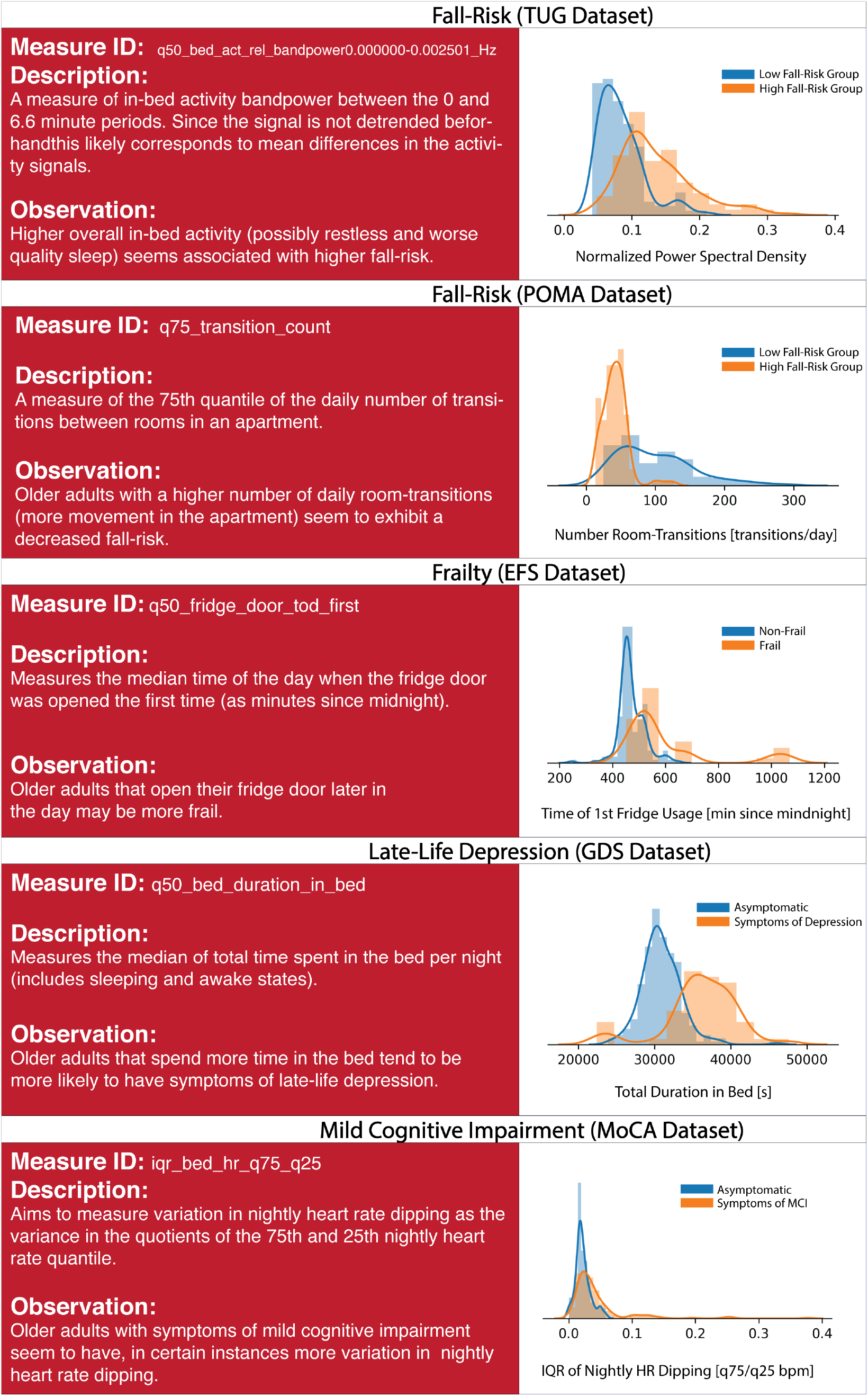
Displays descriptions and density plots of the most important digital measure for each outcome. Across all density plots, blue indicates a positive/neutral outcome, while orange indicates a negative outcome.

**Figure 3:**
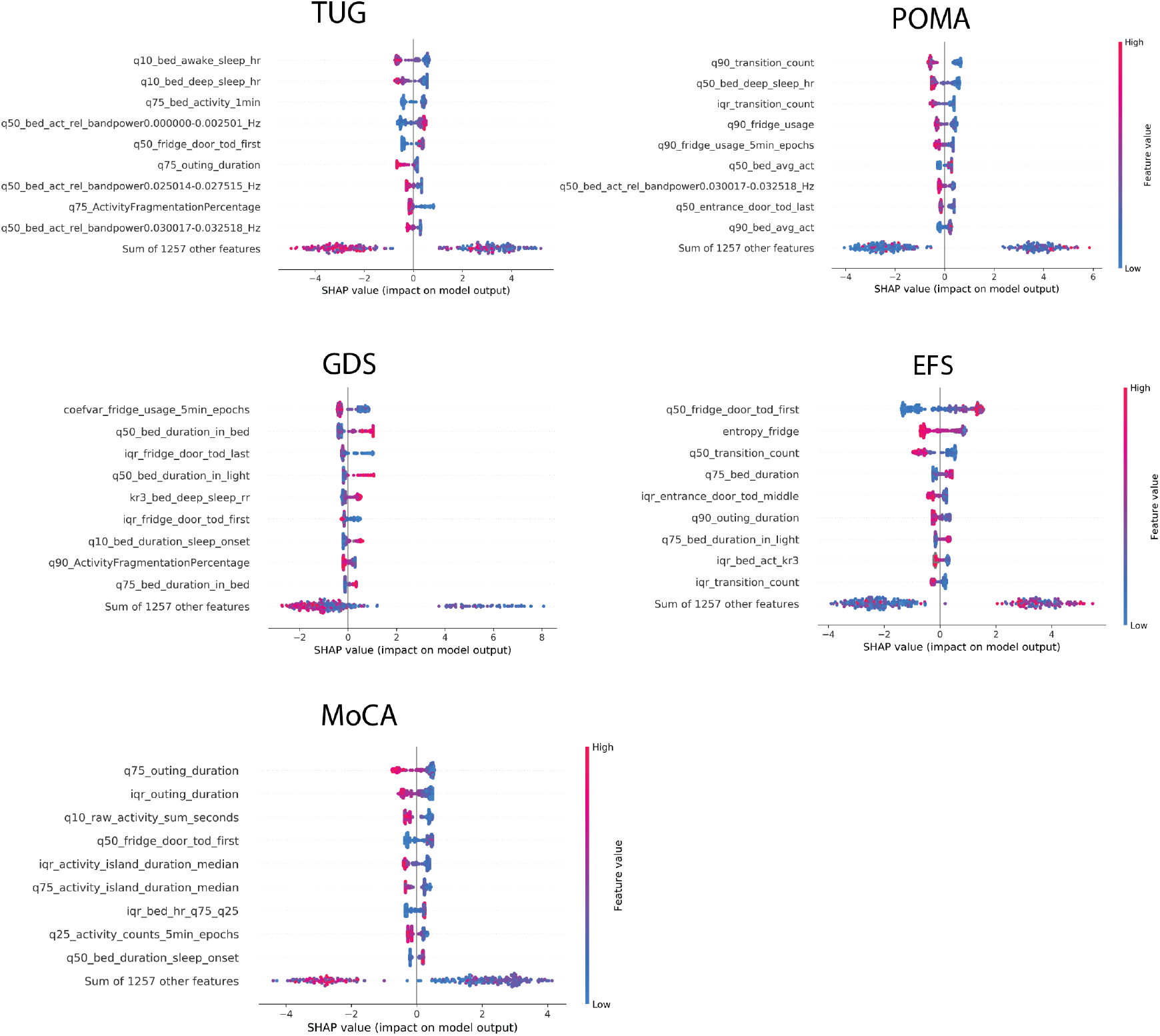
Shows *beeswarm* plots of the 9 most important digital measures based on SHAP values on all outcome datasets: TUG (Timed Up and Go) & POMA (Performance Oriented Mobility Assessment = fall risk, GDS (Geriatric Depression Scale) = late-life depression, EFS (Edmonton Frail Scale) = frailty, MoCA (Montreal Cognitive Assessment) = mild cognitive impairment. Finally, the contributions of the sum of the remaining measures is displayed. Digital measures are ordered according to their importance, from top to bottom. The x-axis represents log odds, where values above zero indicate relevance for a negative outcome. Colouring further shows the direction of this association, where blue indicates lower values of a given measure and red indicates higher respective measure values. Detailed explanations of the individual measure names are given in the supplementary material or on the supplementary website. Note that these plots are based on models trained on the whole respective dataset and are therefore slightly different from the global importances shown in Table C.III, which are based on 100 simulation iterations.

## 3 Discussion

In this work, we introduce a digital behaviorome based on unobtrusive and privacy-preserving contactless sensor technology that is uniquely suitable for use in the population of community-dwelling older adults. We then demonstrate the feasibility of utilising the introduced behaviorome to create machine learning-based digital COAs for common ageing-relevant outcomes, including fall risk, frailty, late-life depression, and mild cognitive impairment. Finally, we determine which individual digital measures were most relevant for a given COA.

While the idea of a sensor-derived behaviorome as a non-exhaustive set of digital measures characterising a person’s behaviour is not entirely new, existing work remains scarce and is not primarily focused on technologies that can be reliably used in the population of community-dwelling older adults. The existing research often concerns the use of wearable devices over brief pre-defined time intervals, which may not be optimal for long-term monitoring (ranging from several years to potentially decades) with older adults; this technology contrasts somewhat with the type of sensor systems that have been successfully used for this purpose. Thus, our introduced digital behaviorome is not only the most detailed (to our knowledge) to date but also derivable by a set of simple, privacy-preserving, and low-cost contactless sensors. Consequently, it is compatible with technologies used in a majority of long-term remote-monitoring projects involving older adults, including the recently established Collaborative Aging Research Using Technology (CART) initiative, which seeks to make ageing-related digital health approaches more accessible to the broader research community [23]. Overall, then, we believe that the presented digital behaviorome has the potential to serve as a baseline set of measures that may be calculated over long time frames (ranging from years to potentially decades), but which could also be supplemented (potentially over shorter time periods) with digital measures based on more specific sensors, such as pillbox door sensors, wearables, or smartphones, depending on the specific needs, circumstances, and conditions.

Our results indicate that a digital behaviorome derived from simple and unobtrusive sensors could be suitable for creating useful ageing-relevant digital COAs. This is reflected by the good discriminative performances across all COAs except for late-life depression, resulting in ROC AUC values of *≥*0.7. Notably, these results are based on a very limited sample size, which makes it highly probable that this is a conservatively low estimate of what could be possible. In all cases, results based on the behaviorome led to higher ROC AUC and PrAUC values than those obtained from only using demographics information such as (see Table 1). These differences were significant in three (TUG, EFS, and MoCA) out of the five outcome datasets, indicating that the behaviorome captures information beyond just simple demographics. It is also notable that adding demographic information to the behaviorome did not result in significantly better performance across outcomes, which may indicate that this type of information is already implicitly present in the behaviorome. While we used COAs here as an example for feasibility purposes, COAs could be very useful for continuously assessing an older adult’s health status with respect to specific outcomes and may allow for the implementation of early preventive interventions. For instance, if an older adult exhibits increased fall risk, it may be reasonable for them to see a fall prevention specialist. Furthermore, COAs such as those shown could aid research into complex ageing-relevant conditions such as cognitive impairment by providing objective information on how such impairment impacts a person’s daily life.

Putting these results into perspective, passive sensor-based fall-risk assessments were shown to yield AUC values in the range of 0.65–0.89 [35]. It should be noted that these values were obtained using wearables by means of accelerometry and predominantly with very few digital measures of gait (and sometimes with accelerometer signal characteristics). Furthermore, none of the studies mentioned in this paper used long-term data, and some were performed under laboratory conditions far removed from real life. Our behaviorome-only results, with ROC AUC values of 0.786 (TUG dataset) and 0.805 (POMA dataset), are thus in line and satisfactory by comparison. Meanwhile, in terms of important digital measures, it is notable that not only physical activity and broadly gait-related measures (such as the number of room transitions) but also sleep and rhythmicity measures, such as activity in bed, bed-exit count, or activity fragmentation, were of major importance in discriminating between participants with high and low fall risk (see Figure 3). Although prospective data would be necessary to form clear conclusions, this may suggest that behavioural data beyond just gait and physical activity may be relevant for fall-risk assessments in older adults. Moreover, results from Piau et al. show that PIR array-based gait speed may help identify future fallers [21]. In this regard, the inclusion of gait-speed information that is more accurate yet still unobtrusive into a behaviorome would likely further increase performance on fall-risk assessments.

In terms of frailty, comparable studies report ROC AUC values between 0.72 and 0.86, based on wearable sensors [36, 37, 38]. These results were primarily obtained on the basis of gait and physical activity measures. Additionally, frailty definitions, study type, and participant characteristics differ quite significantly, so at best this gives a broad idea of what is possible. With a behaviorome-only ROC AUC value of 0.704, our results are on the lower end of this spectrum. However, given the types of sensors on which our behaviorome is based, this seems realistic. Indeed, some of the most important measures related to frailty were related to fridge usage, physical activity (room-transition counts), and sleep duration (see Figure 3), all of which seem plausible as indicators.

For late-life depression, comparable studies are lacking. Several studies have demonstrated the utility of using wearable-based digital measures in assessing general depression [39, 40, 41]. Furthermore, one instance reported on the assessment of late-life depression using PIR-derived information on activities of daily living (ADL) [42]. However, it is unclear whether their methodology prevented data leakage, judging by the unusually high ROC AUC values *≥* 0.95. Our own results, by contrast, show modest performance in assessing late-life depression, with a behaviorome-only ROC AUC value of 0.620. While this may be due to the low number of participants with a GDS score above 5 in our cohorts, it could also indicate some inherent difficulties in measuring this outcome. Many of the most important individual measures for late-life depression assessments are related to sleep duration (see Figure 3), which is known to be associated with depression. More interestingly, variations in fridge usage and behaviour complexity were relevant; however, due to their relatively low discriminative power, further interpretation may not be meaningful.

Regarding the distinction between healthy older adults and those with MCI, recent work has shown ROC AUC values of 0.62–0.80, based on comparable time intervals [43, 32]. These values were achieved with wearable devices but also when using additional modalities such as smartphone and computerised assessments. Our behaviorome-only result, with a ROC AUC value of 0.780, is thus aligned with similar research and shows that good discriminative performance may potentially be achieved through an entirely passive and unobtrusive set of sensors. Further supporting the plausibility of our results, a respectable body of literature shows how individual digital measures based on PIR and door sensors – such as variability in PIR array-based gait speed [44, 45, 20], ADL regularity [13], regularity in physical activity [18], sleep disturbances [46], and outing duration [47] – differed between older adults with MCI and healthy controls. Finally, with regards to MCI, highly important measures include those related to physical activity, such as the number of room-transitions or the total amount of PIR-based activity. Moreover, sleep-related measures such as sleep duration, activity in bed, and variation in in-bed activity were found to be important (see Figure 3). Regarding MCI, however, the most noteworthy finding is the inclusion of various sleep-related heart-rate measures – most importantly, the variation in nightly heart-rate dipping behaviour, where unusually high variation seems indicative of MCI (see Figure 2). This is especially interesting, as it has not been previously reported in connection with digital measures. However, it is known that heart-rate dipping is associated with cardiovascular disease [48] and that cardiovascular risk factors may be involved in cognitive decline [49]. As such, it could be beneficial to further investigate the relationship between nightly heart rate and mild cognitive impairment.

The presented behaviorome is a novel, thoroughly documented, and extensive set of digital measures obtainable with unobtrusive contactless sensor technologies. Sensor technologies appear to be the most successful in real-world use in ageing-related long-term remote-monitoring projects [14]. With growing interest in digital health solutions and remote monitoring, a digital behaviorome may be critical to the future of technology-assisted ageing and may provide value in both healthcare and research settings. As one potential use case, we demonstrate the feasibility of using the introduced behaviorome to create ageing-relevant COAs with real-world data from naturalistic samples of older adults. Our results often align with comparable results obtained from more obtrusive (but potentially more accurate) wearable sensors. Finally, we have found multiple individually interesting digital measures that may merit further investigation. These measures show how a behaviorome may also be of potential use in discovering novel ageing-relevant digital biomarkers. Another notable observation is that, across all outcomes, the sum of the remaining SHAP values – that is, all digital measures except for the 9 most important ones combined – was highly important in explaining model outcomes (see Figure 3). This supports the intuition that an extensive set of digital measures, such as the introduced behaviorome, is likely beneficial in terms of COA performance, as opposed to individual or several digital measures. It should also be mentioned that, by relying entirely on privacy-preserving, unobtrusive, and contactless technologies, large sets of digital measures may be derived without ethical concerns related to burdening subjects with unnecessary sensing modalities. The best practices laid out by Goldsack et at., for instance, discourage efforts towards sensor-symptoms mapping, which is to some degree what a digital behaviorome-based approach is doing [3]. However, since in this case sensor technologies respect privacy and do not add any additional burden, there are scarcely any downsides, as would occur with adding additional wearables or tasks requiring interaction.

Future research should emphasise further analytical and clinical validation of the included digital measures. Moreover, analysing long-term temporal dynamics would be essential, as it would enable the identification of trajectories of certain digital measures or even whole groups of measures. Evaluating trajectories, in turn, could be extremely valuable, as was shown by Akl et al., for instance, who produced impressive results regarding MCI classification based on long-term trajectories of several individual digital measures [20]. In addition, future research could combine a digital behaviorome such as the one we utilised with traditional multi-omics data in a deep phenotyping effort. This may enable a wide range of new research insights into ageing and ageing-related conditions, as it adds a new layer of objective information for characterising phenotypes of health and disease. As a final caveat, a digital behaviorome such as this should never be assumed to be complete or fixed. Future research will add new digital measures while old measures may be merged if they exhibit closely correlated behaviour. As such, in the immediate future, it may be of major value to add more accurate gait-related digital measures to the introduced behaviorome, as these have consistently been shown to be very important across many ageing-relevant health outcomes. Consequently, they are likely to add significant value.

There are four major limitations to this work. First, some of the introduced measures have not been validated beyond the scope of this research (the use or validation of a digital measure in other studies is indicated in the supplementary material). This implies that some digital measures may not quantify what we hypothesise, which could lead to inaccurate interpretations and conclusions. Second, our feasibility demonstration involved a relatively small number of participants. Therefore, these results should be interpreted with caution, although general tendencies are likely to be valid. Third, in the feasibility demonstration, we use a cross-sectional approach that fails to leverage the temporal trajectories of sequentially collected behaviorome slices. Indeed, we strongly believe that data collected over multiple years would be necessary to fully explore the utility of behaviorome-based approaches. Finally, one drawback of simple PIR sensors is that not all digital measures based on this technology can be calculated when more than one person is living in an apartment. This is not a problem for the sleep sensor, and certain PIR sensor-derived measures may be valid nonetheless, but it does warrant consideration.

## 4 Conclusion

In this work, we introduce an unobtrusive digital behaviorome for use in the older adult demographic. Overall, the behaviorome consists of 1268 digital measures derived from 94 hypothesis-driven base measures. All included digital measures are derived from a small set of privacy-preserving contactless sensors that have been successfully used in multiple ageing-related, long-term, remote-monitoring projects around the world. For each measure, we provide a detailed description, background information, and additional real-world data as supplementary online material. While use cases for the introduced behaviorome are diverse, we demonstrate a potential use case by creating multiple machine learning-derived digital COAs and evaluating their discriminative performance. To this end, we show how ageing-relevant outcomes such as fall risk, frailty, and mild cognitive impairment could be assessed through an extensive set of unobtrusively obtained digital measures. Our results with these measures not only show that information from the behaviorome significantly outperformed basic demographic information, but also that behaviorome-based COAs could match the performance reported in comparable studies employing more obtrusive wearable sensors, which may not be optimal for long-term monitoring among older adults. Finally, we highlight the possibility of using a digital behaviorome to potentially discover novel digital biomarkers, using a model explainability approach on the basis of models used to create the aforementioned COAs. The respective results show that the most important digital measures are reasonable, while also revealing two potentially relevant insights. First, while fall risk may be primarily associated with gait and physical activity, it also potentially exhibits strong associations with sleep-related measures. Second, unusually high variation in nocturnal heart-rate dipping was uniquely related to mild cognitive impairment.

## 5 Methods

### 5.1 Minimally-Obtrusive Digital Behaviorome

#### 5.1.1 Sensors

The introduced digital measures are based on three sensor types that have been commonly used in remote-monitoring projects with older adults: PIR sensors, contact door sensors, and a sleep sensor. The PIR motion sensors were placed in the essential rooms of older adults’ apartments. Essential rooms included the living room, bedroom, entrance, bathroom/toilet, and kitchen. The employed PIR sensors sampled with 0.5 Hz, and thus reported activity on or off states every 2 seconds. The reed switch-based door sensors, meanwhile, were placed at the entrance and the refrigerator door. Both PIR and door sensors were part of the DomoCare® (DomoSafety SA, Lausanne, Switzerland) home-monitoring system. Finally, for the sleep sensor, we used an EMFIT QS ferroelectret sensor (Emfit Ltd, Vaajakoski, Finland), which was fixed beneath the mattress at approximately chest height. A summary of these three devices, as well as their respective source data streams, is given in Table 2.

**Table 2:** A summary of the contactless pervasive computing devices used, including the respective data streams they provide.

| Sensor | Device | Location | Device Type | Data Stream | Sampling Rate | Channels |
| --- | --- | --- | --- | --- | --- | --- |
| PIR Motion | DomoCare | Wall/ Ceiling | Contactless | Activity | 0.5 Hz | Entrance<br>Toilet<br>Bedroom<br>Livingroom<br>Kitchen<br>Others |
| Magnetic Door<br>Reed Switches | DomoCare | Door | Contactless | Open/ Close State | Events | Entrance<br>Fridge |
| Bed Sensor | EMFIT QS | Beneath Mattress | Contactless | Heart Rate | 0.25 Hz | N/A |
|  |  |  |  | Respiration Rate | 0.25 Hz | N/A |
|  |  |  |  | Activity | 0.25 Hz | N/A |
|  |  |  |  | Toss & Turns | Events | N/A |
|  |  |  |  | Sleep Phases | 0.25 Hz | Awake<br>REM<br>Deep<br>Light |
|  |  |  |  | Raw Low<br>Frequency Band | 50 Hz | N/A |
|  |  |  |  | Raw High<br>Frequency Band | 100 Hz | N/A |
|  |  |  |  | Bed Exits | Events | N/A |
|  |  |  |  | Summary Statistics | Daily | N/A |

**Table 3:** Participant Characteristics

| Characteristic | Cohort 1 (n=24) | Cohort 2 (n=21) | Pooled (n=45) | Differences (p-value) |
| --- | --- | --- | --- | --- |
| Age (years) | 88±7 | 86±7 | 87±7 | 0.50 |
| Sex female (%) | 79 | 52 | 67 | 0.06 |
| TUG | 14.0±10.0 | 15.3±4.8 | 14.5±8.2 | 0.58 |
| POMA | 21.4±7.4 | 24.1±3.3 | 22.5±6.1 | 0.13 |
| EFS | 4.6±3.6 | 5.0±1.7 | 4.7±2.9 | 0.65 |
| GDS | 2.3±2.3 | 4.2±3.2 | 3.1±2.9 | 0.04 |
| MoCA | 20.5±5.1 | 20.6±3.7 | 20.5±4.7 | 0.95 |

#### 5.1.2 Creation of Digital Measures

The creation of the digital measures was mostly hypothesis-driven or based on measures from the existing literature. The majority of measures were calculated on a daily or nightly basis (for instance, daily total activity, daily outing duration, or average heart rate during a night). Subsequently, we calculated derivates of those measures by means of descriptive statistics over non-overlapping bi-weekly segments.While bi-weekly segments may be somewhat arbitrary, two weeks is a sufficient period to capture variation in behaviour that often follows daily or weekly cycles while still being short enough to capture temporally limited behaviours. Additionally, it serves to increase the number of data points (data augmentation) and facilitates the process of working with sensor recordings of various lengths or with data gaps. For cases of certain behavioural or rhythmicity measures, such as Cosinor regression-based measures, raw data from the whole bi-weekly segment was used directly. To avoid the inclusion of measures with insufficient data, we set a minimum number of 10 days for which raw source data was available throughout a given bi-weekly segment; otherwise, the measure was set as missing. The criteria for including a day’s worth of data for each sensor type are explained in detail in the supplementary material 1.

For all daily or sub-daily base measures, derivates based on summary statistics were calculated over the bi-weekly segments. Summary statistics include various quantiles, denoted as q*n* (e.g. q10), the interquartile range (iqr), the mean, median (=q50), coefficient of variation (coefvar), and robust measures of kurtosis and skewness (kr3 and sk3, respectively), following the naming convention proposed by Kim and White [50]. Figure 4 summarises this workflow visually. Eventually, this left 1268 dimensional vectors (one per bi-weekly segment). Of those, 223 dimensions are related to PIR and door sensors, while the remaining 1045 are based on sleep-sensor data. Detailed information regarding the exact calculation of each measure, as well as individual distributions across our cohort, can be found at *online*.

**Figure 4:**
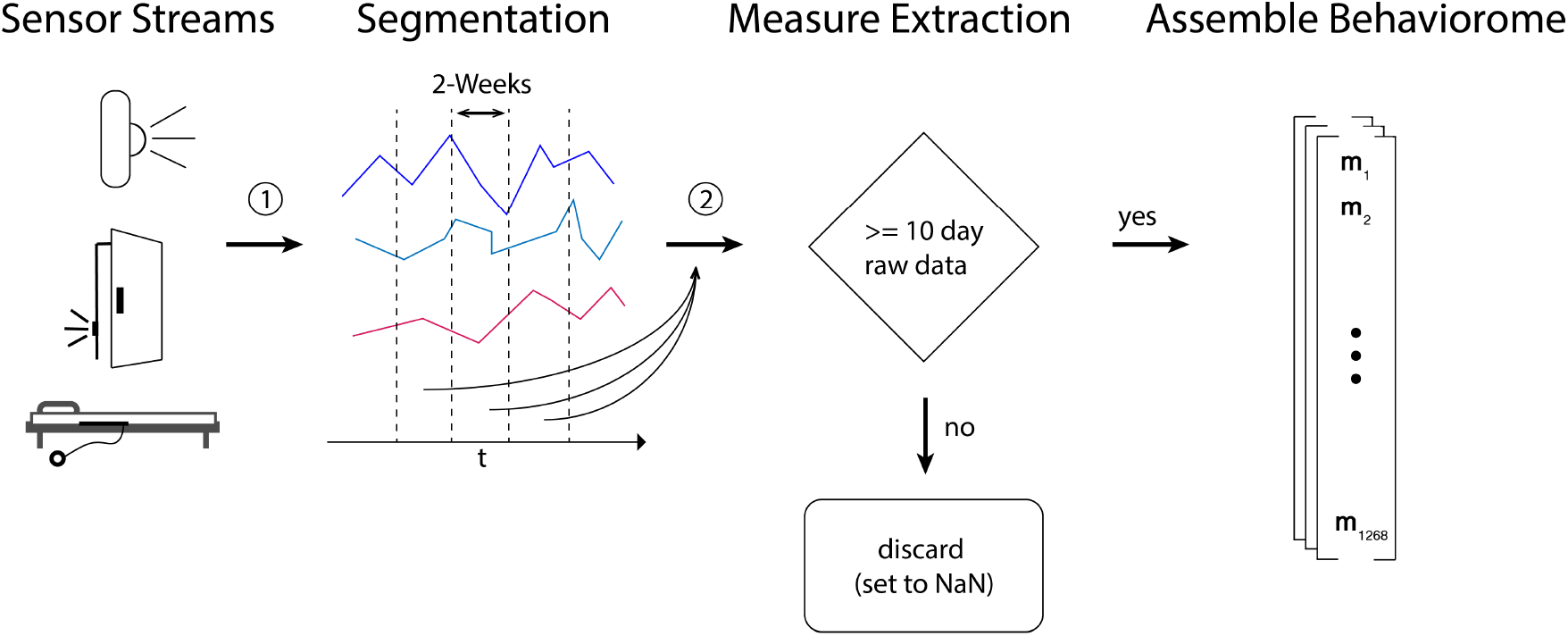
Shows a broad summary of how digital measures were calculated, starting with raw sensor data from PIR sensors, door sensors, and bed sensors. Raw data streams were first segmented into non-overlapping bi-weekly segments. Then, for each bi-weekly segment, digital measures were calculated. If, for a given measure, less than 10 days of data were present, the measure was encoded as missing, which eventually left 1268 dimensional vectors – one per bi-weekly segment.

#### 5.1.3 Example Visualisation

We provide an example of averaged behavioromes with respect to MCI. These visualisations were created by first averaging the behavioromes on a per-participant basis, followed by z-normalisation. After that, the behavioromes were split into the positive and negative outcome groups and averaged once more. Finally, heatmaps for both conditions were created. As a result, the values of individual digital measures *>* 0 indicate above-average values, while those *<* 0 indicate below-average values.

### 5.2 Machine Learning based Digital Outcome Assessments

#### 5.2.1 Participants

To test the feasibility of using the previously described behaviorome to create ageing-relevant COAs, we used real-world remote-monitoring data from two cohorts of older Swiss adults (pooled age years=87 *±* 7; sex 67% [30/45] female). The original studies were both pilots designed to assess novel computing technologies for ageing-in-place scenarios in the German- and French-speaking cantons of Switzerland [51, 52]. They were conducted between 2017 and 2018 and monitored participants over one year with a set of pervasive computing devices and clinical assessments [51, 52]. The inclusion criteria between cohorts were similar in the sense that both aimed to recruit a natural sample of community-dwelling older adults (aged *≥*70 years) who lived alone and without pets. On the other hand, the exclusion criteria between cohorts differed. For cohort 1, the only exclusion criterion was an unwillingness to comply with the study protocol. But, for cohort 2, the exclusion criteria were as follows: (1) severe cognitive impairment rendering the individual unable to follow study protocol (clock-drawing score *≥*4); (2) skin problems such as irritations, itching, or serious redness; (3) undergoing dialysis; (4) unwillingness to comply with the study protocol; (5) an inability to understand the study aim; or (6) hospitalisation planned within a short period of time [51]. Both studies were conducted based on principles declared in the Declaration of Helsinki and approved by the Ethics Committees of the cantons of Bern and Vaud (KEK-ID: 2016-00406 and CER-VD ID: 2016-00762, respectively). All subjects signed and returned informed consent forms before participating in the study. Detailed participant characteristics and cohort differences are shown in the Appendix 3. The differences between cohorts were statistically examined on the basis of unpaired, two-sided, two-sample t-tests (*α* = 0.05). In every analysis involving participant data, all participants with any available data (depending on sensor data and the availability of clinical assessments) were included; this also applies to participants that dropped out of the studies.

### 5.3 Clinical Assessments

Participants in both cohorts were subject to an overlapping set of standardised clinical assessments. These include the following six assessments: (1) the Timed Up and Go Test (TUG), which is often used in geriatrics to assess fall risk [53]; (2) the Tinneti Performance-Oriented Mobility Assessment (POMA), which, as with the TUG, also measures balance and gait characteristics that are often indicators for elevated fall risk among older adults [54]; (3) the Edmonton Frail Scale (EFS), a frequently used measure of frailty among older adults [55]; (4) the short version (15-item) of the Geriatric Depression Scale (GDS), a commonly used late-life depression screening tool [56]; (5) the Montreal Cognitive Assessment (MoCA), which measures cognitive function and is often used as a brief screening tool for the detection of MCI in older adults [57]. In each cohort, these assessments were planned to be conducted at least once during the one-year study duration. Detailed assessment intervals are summarised in Table 4.

**Table 4:**
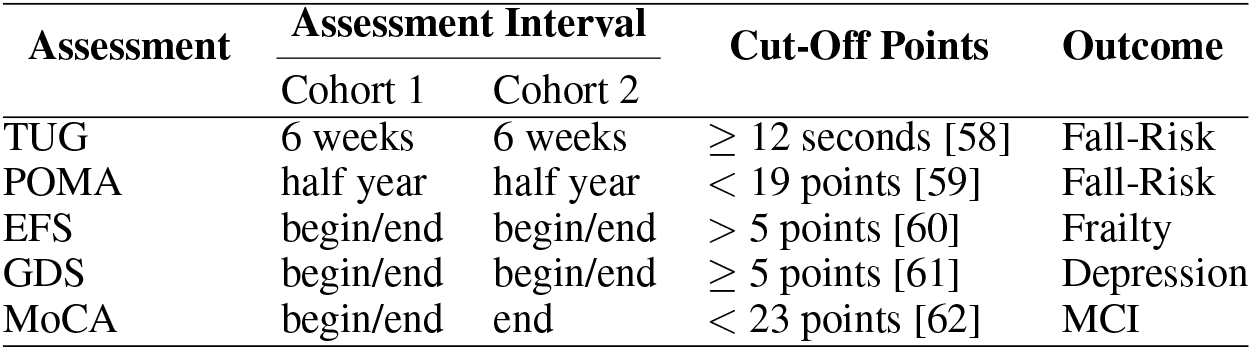
Overview of clinical assessments used to evaluate the health status of study participants, including the measurement interval and cut-off points used to divide participants into groups with positive/neutral or negative health outcomes.

### 5.4 Dataset Creation

To evaluate the potential for creating COAs that may help differentiate between positive and negative ageing-relevant health outcomes based on the proposed behaviorome, we categorise participants into one of the two categories for each clinical assessment. This was done on the basis of validated cut-offs for each assessment in 5.3. The respective cut-off values for the negative groups are shown in Table 4. Next, we calculated the digital measures as presented in 5.1.2 for all participants. For each assessment, we then combined the positive/negative labels with the bi-weekly segments of a given participant. If multiple records of the same clinical assessments were obtained throughout the study, we assigned the target label corresponding to the assessment closest in time. After this procedure, we obtained one dataset per assessment.

Note that measures derived from PIR and door sensors stem from one sensor system (meaning that technical failure usually affect both sensor types, except for instances were an individual sensor unit failed, which happened rarely), while sleep stems from another sensor; thus, for a bi-weekly segment to be valid, at least 30% of measures from both sensor systems must be valid. This led to a significant reduction in the number of bi-weekly segments, as a large number of sleep sensor data were missing due to technical issues, as has been discussed in prior work [16]. These two issues — lacking sensor data from both PIR/door and bed sensors and the unavailability of respective assessments — are responsible for the generally lower numbers of participants who could be included in this analysis (the exact numbers with regards to each assessment are given in Table 5). In Figure 5, we present the high-level flowchart of dataset creation.

**Table 5:** Dataset Characteristics

| Dataset | Participants Total | Participants with Negative Outcome | Number Bi-Weekly Segments |
| --- | --- | --- | --- |
| TUG | 28 | 14 | 277 |
| POMA | 28 | 10 | 277 |
| EFS | 28 | 10 | 277 |
| GDS | 28 | 7 | 277 |
| MoCA | 25 | 16 | 260 |

**Figure 5:**
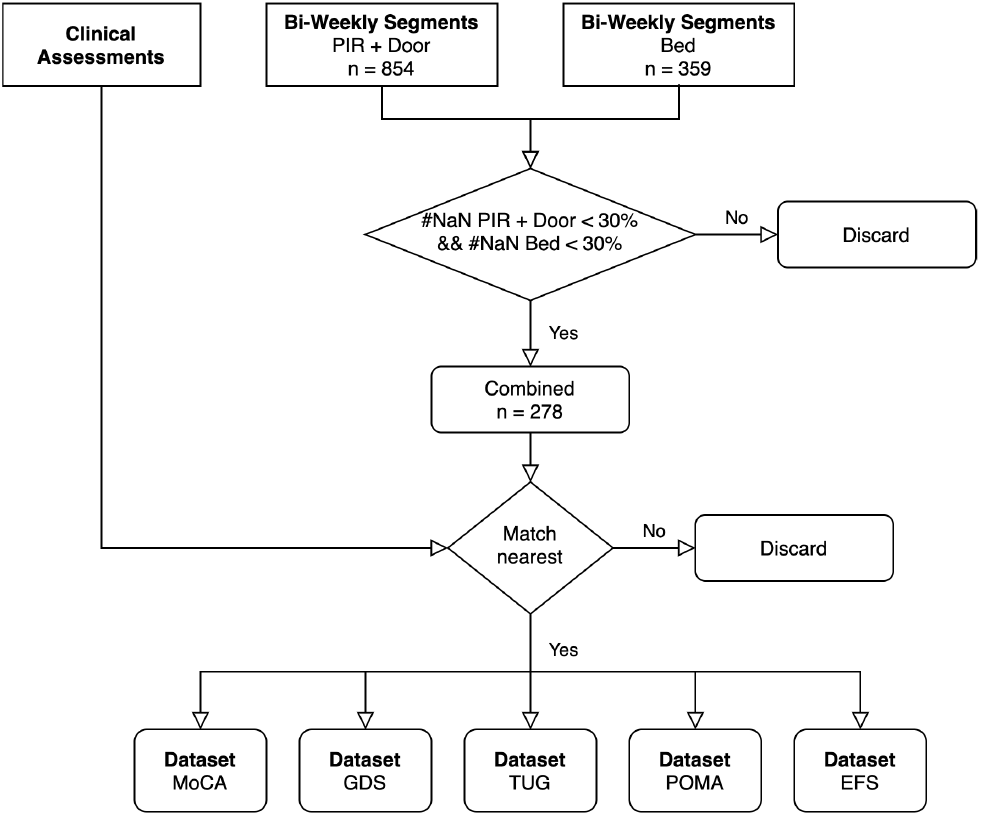
Highlights the workflow of creating datasets, subsequently used for the creation of digital clinical outcome assessments. First, digital measures were separately calculated for the PIR + door and bed sensors and segmented into non-overlapping, bi-weekly segments. After that, the measures from bi-weekly segments, where the percentage of missing digital measures from either sensor system was *<* 30%, were combined. Next, the clinical assessments from each participant were matched with the respective bi-weekly digital measure vectors to combine 5 datasets — one for each assessment.

### 5.5 Machine Learning Analysis

A small forenote aimed at a more technically oriented audience: what we call digital measures throughout this work can be seen as synonymous to the more abstract and general term “features”. To evaluate COAs based on the behaviorome, we largely followed the approach set out by Chen et al., albeit with some minor changes [32]. As such, we use the gradient boosting-based XGBoost algorithm [63] as a classifier, since it generally performs impressively on tabular data, tends to deal reasonably well with high-dimensional feature spaces (even in *p >> n*-type scenarios, as here), and can inherently deal with missing values, all of which means it is close to being the gold standard for this kind of application [32, 64, 39]. Furthermore, gradient boosting-based tree approaches tend to be more easily explainable than modern neural network approaches such as convolutional neural networks, while also retaining high accuracy, especially on tabular data structures [34]. To better account for stochasticity in participant selection, we further adapted the simulation strategy of Chen et al. [32], in which 70% of participants were repeatedly drawn from the entire participant pool to form a training set, while the remaining 30% were used as a test set (the splits are stratified for the respective clinical assessment labels). This procedure was repeated for 100 iterations. Note, that this way each new draw represents a shuffling of the dataset without introducing data leakage between training and test splits. Throughout each iteration, hyperparameters were first optimised within the training split by means of stratified 3-fold cross-validation coupled with random search (consisting of 50 search trials). For more detailed explanation of this strategy, we refer to the original article by Chen et al., where it is demonstrated in detail [32]. Eventually, for each iteration, we calculated the Area Under the Receiver-Operating Curve (ROC AUC) and the Area Under the Precision Recall Curve (PrAUC) on the test set, where multiple bi-weekly segments from a single participant were combined into one score by averaging their predictions (soft voting), as was done in [32]. Likewise, if multiple assessment results were available, they were first averaged, which should have also reduced some of the inherent noise; these results were then dichotomised on the basis of the previously introduced cut-off points (see Table 4) to yield a single label per participant. We removed three digital measures (Measure IDs: *iqr_entrance_door_tod_first, q50_entrance_door_tod_first*, and *q50_fridge_door_tod_middle*) from the full set for this portion of the analysis, as they were biased towards identifying one of the two cohorts (to account for further less-obvious biases in this regard, we included cohort information in the demographics). Note that the PrAUC is sensitive to label distribution, which means it only lends itself to comparisons within the same dataset. For each assessment, we ran three different scenarios, one with only demographic information (age, sex, and cohort membership) as baseline, one with only the behaviorome, and one with both the behaviorome and demographic information combined. Differences between these scenarios were deemed statistically significant if the 95% CIs of two conditions do not overlap. Model hyperparameter ranges were given in the supplementary material. We used the original Python (version 3.6) implementation of XGBoost (version 1.3.3). Model training was performed on UBELIX (http://www.id.unibe.ch/hpc), the HPC cluster at the University of Bern.

### 5.6 Identifying Digital Measures of Interest

To better understand the role of individual digital measures in machine learning-based COAs, we used SHapley Additive exPlanations (SHAP), a game-theoretic approach for explaining complex machine learning models. With this approach, exact solutions can be found in the case of tree-based models [33, 34]. SHAP values have been used fairly extensively in recent biomedical applications [65, 32, 66, 67]. For each of the assessments, we provide overall global SHAP values across all 100 simulations. That is, we give the mean absolute value of the SHAP values for a given digital measure *m* in a single simulation, summed up over all simulations

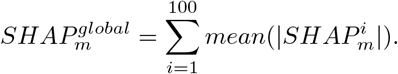

While global SHAP values reveal the overall importance of a given digital measure, they do not say anything about the direction in which the digital measure influences a model. Therefore, we additionally calculate beeswarm plots of the SHAP values. These are based on a model trained over the entire respective dataset, with manually set hyperparameters (reported in the supplementary material). SHAP values were calculated using Python (version 3.6) with the *shap* package (version 0.39.0).

## Supporting information

Detailed descriptions of each individual base measure.

Summary table of presented digital measures.

## Data Availability

Original data used in this manuscript may be obtained upon request but will require ethical approval from the responsible authorities.
Some aggregated data related to the manuscript is available online.

https://narayanschuetz.github.io/digital-behaviorome/

## Appendix

### 1 Individual Day and Night Data Inclusion Criteria

#### 1.1 PIR and Door Sensors

Sensors associated with the DomoCare system (PIR and door) were marked invalid based on whole days. A day is deemed invalid, if less than 300s of raw PIR activity was present for the whole apartment. We found that in general, days with less activity were corresponding to technical issues or extremely long outings (such as related to holidays or hospitalizations). In Figure A.1a, this cut-off value is indicated. It is visible, how there is a second modality in the distribution appearing close to zero, distinct from the main modality, referring to the normal range of daily activity sums.

#### 1.2 Bed Sensor

For night inclusion, we adopt the same procedure as used in previous work [16], displayed in A.1b. Nights without any data were marked as invalid beforehand.

**Figure A.1:**
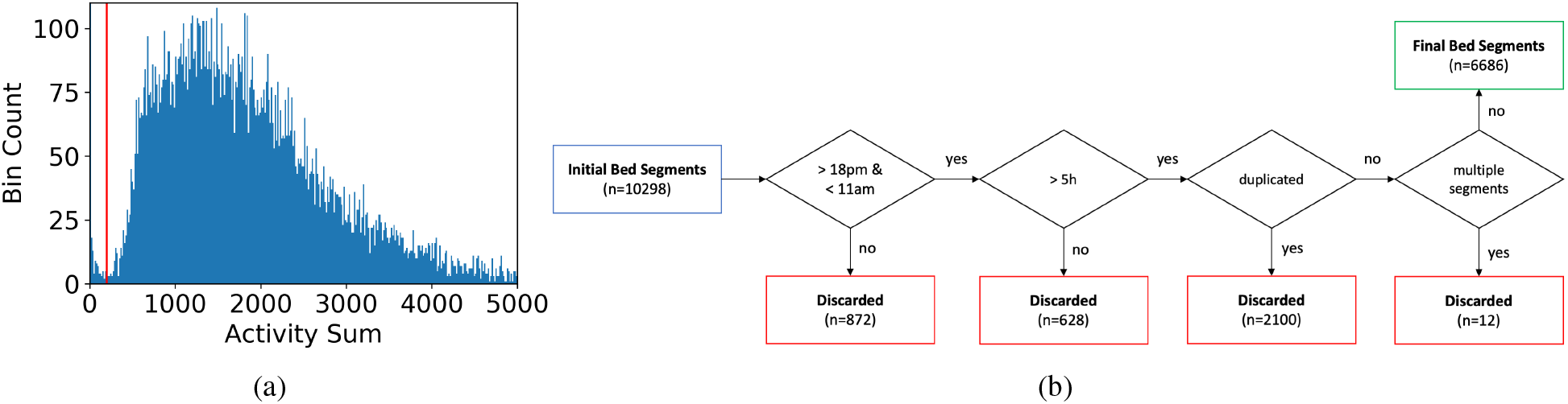
a) Shows raw PIR sensor system activity distribution for individual days, across all participants of both cohorts. The red vertical line denotes the cut-off, corresponding to the minimal activity in seconds for a day to be valid. Days below this value likely correspond to non-normal days, such as days with technical issues or days, where participants were away from home for almost the whole day. b) Depicts the inclusion criteria for nights to be valid based on sleep sensor data. If no data is present at all, the whole night is marked invalid. (the depicted flowchart is based on our original article in https://mhealth.jmir.org/2021/6/e24666).

### 2 Hyperparameter Search Space

In Table B.I we list the hyperparameter search space that was optimized with 3-fold cross-validation within each training split. Remaining hyperparameters were left at default values.

**Table B.I:**
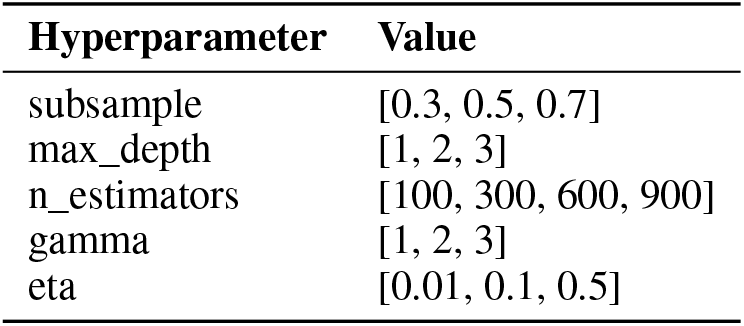
Hyperparameters of models used to produce beeswarm SHAP plots.

### 3 Beeswarm SHAP Values Plot Hyperparameters

In table C.II we list the XGBoost hyperparameters, used to train the models the SHAP *beeswarm* plots are based upon. Note, these hyperparameters are based on manual fine tuning and resulted in good performance across all outcome datasets. Hyperparameters not mentioned were left at default values.

**Table C.II:**
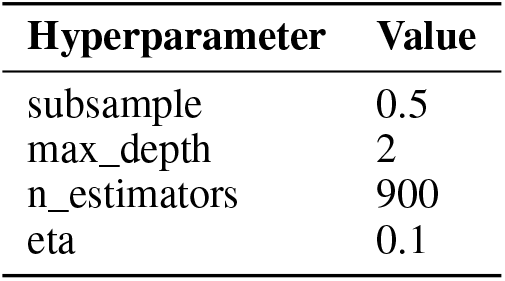
Hyperparameters of models used to produce beeswarm SHAP plots.

| **Hyperparameter** | **Value** |
| --- | --- |
| subsample | 0.5 |
| max_depth | 2 |
| n_estimators | 900 |
| eta | 0.1 |

**Table C.III:** Displays digital measures sorted on the basis of highest global SHAP importances.

| Digital Measure ID | Global SHAP Value |
| --- | --- |
| *TUG Dataset* |  |
| q50_bed_act_rel_bandpower0.000000-0.002501_Hz | 55.755 |
| q50_fridge_door_tod_first | 23.808 |
| q50_bed_act_rel_bandpower0.025014-0.027515_Hz | 21.877 |
| q75_bed_duration | 15.889 |
| q25_bed_nocturia_count | 10.104 |
| q90_bed_bedexit_count | 9.163 |
| q50_bed_act_rel_bandpower0.030017-0.032518_Hz | 8.994 |
| q75_outing_duration | 7.800 |
| q90_ActivityFragmentationPercentage | 7.800 |
| q90_bed_duration_in_sleep | 7.435 |
| *POMA Dataset* |  |
| q75_transition_count | 37.061 |
| q50_bed_act_rel_bandpower0.025014-0.027515_Hz | 23.863 |
| q75_fridge_usage_5min_epochs | 21.461 |
| q90_ActivityFragmentationPercentage | 17.651 |
| q90_transition_count | 13.879 |
| q90_fridge_usage | 13.645 |
| iqr_transition_count | 12.639 |
| lfhf | 11.947 |
| q75_fridge_usage | 11.107 |
| q50_bed_deep_sleep_hr | 10.599 |
| *EFS Dataset* |  |
| q50_fridge_door_tod_first | 55.915 |
| entropy_fridge | 49.930 |
| q50_transition_count | 19.039 |
| q75_transition_count | 18.332 |
| q90_bed_duration | 15.546 |
| q10_transition_count | 13.577 |
| q75_bed_duration | 13.066 |
| q90_fridge_usage_5min_epochs | 11.982 |
| iqr_outing_duration | 11.275 |
| q50_fridge_door_tod_last | 10.826 |
| *GDS Dataset* |  |
| q50_bed_duration_in_bed | 44.054 |
| q75_bed_duration_in_bed | 25.409 |
| q50_bed_duration_in_light | 23.934 |
| iqr_fridge_door_tod_last | 22.334 |
| iqr_fridge_door_tod_first | 21.360 |
| reconstruction_error | 18.220 |
| q75_bed_awake_sleep_hr | 14.743 |
| coefvar_fridge_usage_5min_epochs | 14.063 |
| q50_bed_duration | 13.913 |
| q75_bed_duration_in_light | 12.611 |
| *MoCA Dataset* |  |
| iqr_bed_hr_q75_q25 | 31.798 |
| q50_bed_act_rel_bandpower0.000000-0.002501_Hz | 18.528 |
| q50_transition_count | 16.585 |
| q90_bed_duration | 14.927 |
| q90_bed_activity_1min | 14.562 |
| q50_bed_hr_rel_bandpower0.007504-0.010006_Hz | 11.331 |
| q75_raw_activity_sum_seconds | 9.295 |
| q90_bed_bedexit_count | 8.658 |
| q50_bed_hr_rel_bandpower0.005003-0.007504_Hz | 8.461 |
| coefvar_bed_activity_1min | 8.261 |

## Footnotes

1 https://narayanschuetz.github.io/digital-behaviorome/SummaryTable.xlsx

2 ^2^https://narayanschuetz.github.io/digital-behaviorome

