## Supplementary material for "An aging focused unobtrusive and Privacy-Preserving Digital Behaviorome": Detailed descriptions of each individual base measure.

### DESCRIPTION OF AGING FOCUSED UNOBTRUSIVE DIGITAL MEASURES

TECHNICAL REPORT

**Narayan Schütz**
ARTORG Center for Biomedical Engineering Research
University of Bern
Bern, Switzerland


**Samuel E.J. Knobel**
ARTORG Center for Biomedical Engineering Research
University of Bern
Bern, Switzerland


**Angela A. Botros**
ARTORG Center for Biomedical Engineering Research
University of Bern
Bern, Switzerland


**Michael Single**
ARTORG Center for Biomedical Engineering Research
University of Bern
Bern, Switzerland


**Tobias Nef**
ARTORG Center for Biomedical Engineering Research
University of Bern
Bern, Switzerland


December 23, 2021

#### ABSTRACT

This is a technical protocol, introducing a digital behaviorome, consisting of an extensive set of 1268 hypothesis driven digital measures of health, obtainable through a small set of minimally-obtrusive contactless sensors. The underlying sensor technologies have been used for more than a decade in ageing related long-term, remote-monitoring projects and been shown to be practical and accepted by older adults across continents. The digital measures are based on 94 base measures, where for each we provide background and hypothesis, as well as references to previous use. In addition, a description detailing how to calculate each measure is provided, including pseudo-code and relevant formulas - where appropriate.

***Keywords*** Digital Measures of Health · Digital Biomarkers · Health Monitoring

### 1 Introduction

This is a supplementary article, detailing a minimally-obtrusive set of hypothesis driven digital measures. All presented measures can be calculated based on a bed sensor, door contact sensors and passive infrared motion sensors placed in essential rooms of a subject. Essential rooms include: entrance, living room, bedroom, bathroom/toilet, kitchen. Doors equipped with sensors include entrance doors and fridge door. For more details with regards to the sensors we refer to the original publication. Here we give a detailed description of 94 digital base measures, based upon which we subsequently derived the full set of 1268 digital measures, making up the introduced behaviorome. We give background information about each measure, for instance where and how they have been previously used, if applicable. Additionally, exact details on how to calculate the measures are provided, including pseudo-code, where applicable.

To ease in interpretability of the shown measures, we categorized them into the following broad categories, where in some cases multiple catagories apply: Physical Activity, Behavior Complexity, Rhytmicity, Sleep, Sleep Complexity, Social Activities, and Activitiesof Daily Living. The *Measure Ids* of all digital measures as used for the programmatic analysis are given under the respective section of each base measure. This includes derived measures based on summary statistics or other parametrizations, such as different frequency bands. The majority of introduced measures are based on a daily or nightly scale, meaning they are derived from a single day or night worth of data. A full summary of the whole behaviorome, including over what time-scale it was calculated, is given in an additional supplementary Excel table (`https://narayanschuetz.github.io/digital-behaviorome/SummaryTable.xlsx`).

The information presented here is also available on a supplementary website with additional information and interactive visualizations (`https://narayanschuetz.github.io/digital-behaviorome`).

### 2 Digital Measures

#### 2.1 Passive Infrared and Door Sensor Derived Measures

##### 2.1.1 Activity Fragmentation

***Summary***: Activity fragmentation measures how fragmented a person's in-home activity is. Measured as probability.

***Measure Ids***

1. *activity_fragmentation*

***Type***

1. *Rhythmicity*

2. *Behavior Complexity*

**2.1.1.1 Background and Hypothesis** Activity fragmentation measures how fragmented a person's in-home activity is. The measure is inspired by the notion of sleep fragmentation, as for instance used in [Lim et al., 2013]. Broadly speaking, activity fragmentation is calculated as the probability $Pr(Inactive|Active)$ of transitioning from an active state to an inactive activity state in a state space model with the two states $Active$ and $Inactive$ - representing whether activity occurred in a certain time-epoch. The hypothesis here is that a person with highly continuous bouts of activity will have a lower transition probability, while one with highly irregular and rather erratic activities will have a higher transition probability. Conditions such as cognitive decline may lead to more fragmented activities as daily activities may be less planned for. On the other hand, problems with mobility or depression may lead to less fragmented activities as a person tries to actively minimize activities where possible.

**2.1.1.2 Description** The activity fragmentation is calculated by computing non-overlapping 5 minute epochs from the passive infrared (PIR) sensor activities signals. These epochs are then further transformed into states active (if any PIR sensor activation occurred in a given epoch) or inactive (if no PIR sensor activation occurred in a given epoch). Based on these states, the probability $Pr(Inactive|Active)$ of transitioning form an active to an inactive state is estimated. It should be noted, that missing values due to outing behavior is less problematic here as $Pr(Inactive|Active)$ does not factor in inactivity per-se but rather the state transitions. A detailed description is given in algorithm 1.

**Algorithm 1:** Calculate Activity Fragmentation

```
function calculate_activity_fragmentation (M);
Input: Matrix M containing PIR sensor firings with columns for time and duration of each activation
Output: Vector of daily activity fragmentation estimates
Initialize list activity_fragmentations;
MIN_ACTIVITY = 300;
days = partition_into_days(M);
foreach day ∈ days do
    motion = calculate_motion_sum(day);
    if motion > MIN_ACTIVITY then
        motion_epochs = calculate_5minute_epochs(day);
        previous_state = 0;
        active_to_inactive = 0;
        active_to_active = 0;
        foreach motion_epoch ∈ motion_epochs do
            state = 0;
            if sum(motion_epoch) > 0 then
                state = 1;
            if state == 1 && previous_state == 1 then
                active_to_active++;
            if state == 0 && previous_state == 1 then
                active_to_inactive++;
            previous_state = state;
        end
        transition_probability = active_to_inactive / (active_to_inactive + active_to_active);
        activity_fragmentations.insert(motion);
end
return activity_fragmentations;
```

###### 2.1.2 Cosinor Single Component Acrophase

***Summary***: The activity acrophase is a commonly calculated measure in chronobiology often derived from actigraphy data. Measured as radians [rad].

***Measure Ids***

1. *one_component_acrophase*

***Type***

1. *Rhythmicity*

**2.1.2.1 Background and Hypothesis** The activity acrophase is a commonly calculated measure in chronobiology often derived from actigraphy data. It describes the phase shift angle of the activity peak on the basis of a fitted cosinusoidal curve with a - usually - 24h period [Cornelissen, 2014]. In simpler terms, it describes the time of day when a person is most active. Acrophase has been described as being associated with a variety of health conditions, as for instance Alzheimer's disease and mood disorders [Li et al., 2020, Smagula, 2016]. Since a large part of activity happens in-home in older adults, in-home activity can be a reasonable proxy of activity, even more so, if we are interested in activity patterns and not primarily the raw numbers (which may require calibration to be comparable across different apartments) [Schütz et al., 2019, 2020]. We therefore estimate cosinusoidal curve parameters based on PIR sensor in-home activity patterns.

**2.1.2.2 Description** The single component trigonometric regression model used is given as

$$y(t) = A_1 \sin\left(\frac{t}{24} 2\pi\right) + A_2 \cos\left(\frac{t}{24} 2\pi\right) + M + e(t), \tag{1}$$

where $A_{1,2}$ and $M$ are model parameters to be learned and $e$ represents an error term, accounting for unexplained variation [Moškon, 2020]. The best fit can be found by minimizing the residual sum of squares, as with regular linear regression. Following [Moškon, 2020], the acrophase $\phi_a$ can be calculated as:

$$\phi_a = \begin{cases} -\arctan\left(\left|\frac{A_1}{A_2}\right|\right) & A_1 > 0, A_2 > 0, \\ -\pi + \arctan\left(\left|\frac{A_1}{A_2}\right|\right) & A_1 > 0, A_2 < 0, \\ -2\pi + \arctan\left(\left|\frac{A_1}{A_2}\right|\right) & A_1 < 0, A_2 > 0, \\ -\pi - \arctan\left(\left|\frac{A_1}{A_2}\right|\right) & A_1 < 0, A_2 < 0. \end{cases}$$

To calculate the activity acrophase from PIR sensor activity we first transform the raw PIR activity signals of each monitored day to non-overlapping hourly bins (we observed that lower time resolutions lead to more stochasticity). In a second step, this hourly activity data is used to fit the trigonometric function (Equation (1)), from which $\phi_a$ can be calculated. Hours that overlap with outings were imputed by their the respective temporal mean without outing, multiplied by 1.4. For the cosinor curve fitting, we used the python implementation CosinorPi [Moškon, 2020]. Algorithm 2 describes the procedure use to arrive at all cosinor based parameters.

**Algorithm 2:** Calculate Cosinor Coefficients

```
function calculate_cosinor_coefs (M, D, E);
Input: Matrix M containing PIR sensor firings with columns for time and duration of each activation, matrix D
  containing entrance door events with occurrence times and matrix E containing sensor events from other in-home
  sensors that mark presence (fridge sensor, bed sensor).
Output: Estimate of cosinor parameters: mesor, acrophase, n_components, amplitude
Initialize list activity_bins;
MIN_ACTIVITY = 300;
days = partition_into_days(M);
foreach day ∈ days do
    motion = calculate_motion_sum(day);
    if motion > MIN_ACTIVITY then
        outings = calculate_outings(day, D, E);
        outing_estimate = calculate_temporal_activity_means(days, D);
        hourly_activity_bins = calculate_hourly_activity_bins(day);
        for i ∈ 0:length(hourly_activity_bins)-1 do
            if hourly_activity_bins[i].overlaps_with(outings) then
                hourly_activity_bins[i] = calc_mean_wo_outing(i, days, D) * 1.4;
        end
        activity_bins.insert(hourly_activity_bins);
end
cosinor_coefficients = fit_cosinor(activity_bins);
cosinor_coefficients["normalized_amplitude"] = cosinor_coefficients["amplitude"] /
  standard_deviation(activity_bins);
cosinor_coefficients["normalized_mesor"] = cosinor_coefficients["mesor"] / standard_deviation(activity_bins);
return cosinor_coefficients;
```

###### 2.1.3 Cosinor Multi Component Acrophase

***Summary***: Acrophase based on PIR activity, allowing multiple components in the Cosinor regression formulation. Measured as radians [rad].

***Measure Ids***

1. *multi_component_acrophase*

***Type***

1. *Rhythmicity*

**2.1.3.1 Background and Hypothesis** The multi component acrophase is basically the same as the single component one described in 2.1.2, except here the cosinor regression is composed of potentially multiple terms, allowing to fit more complex activity rhythms.

**2.1.3.2 Description** Similar to its single component variant, the model used for curve fitting is defined as

$$y(t) = \sum_{i=1}^{N} \left( A_{i,1} \, \sin\left(\frac{t}{24/i} \, 2\pi\right) + A_{i,2} \, \cos\left(\frac{t}{24/i} \, 2\pi\right)\right) + M + e(t). \tag{2}$$

The optimal number of components was chosen based on the extra sum-of-squares F-test, taking into account the increased degrees of freedom when adding more components [Moškon, 2020]. We restricted $N \in \{1, ..., 6\}$, as more parameters would likely lead to overfitting behavior, given we estimate parameters on 10 days of PIR activity data. Calculating $\phi_a$ here is not analytically possible and has to be estimated from the fitted curve [Moškon, 2020]. A detailed description is given in algorithm 2. As with the single component variant, we used the python implementation CosinorPy for multi-component curve fitting [Moškon, 2020].

##### 2.1.4 Cosinor Single Component Mesor

***Summary***: Rhythmicity adjusted activity mean based on a single component Cosinor model. Measured as PIR Activity [s].

***Measure Ids***

1. *one_component_mesor*
2. *one_component_mesor_normalized*

***Type***

1. *Rhythmicity*

**2.1.4.1 Background and Hypothesis** The activity mesor $M$ is a rhythmicity adjusted activity mean, obtainable by means of cosinor curve fitting. It can be seen as a measure of the overall activity, but is due to limited meaningfulness rarely used [Paudel et al., 2010, Neikrug et al., 2020]. There are limited studies that found lower mesor in depressed compared to healthy individuals [Minaeva et al., 2020]. Overall a higher value here refers to higher physical activity levels.

**2.1.4.2 Description** The activity mesor $M$ in equation (1) is directly estimated by fitting the respective model. The procedure to arrive at the mesor is the same as for the other cosinor derived coefficients, see algorithm 2 for details.

##### 2.1.5 Cosinor Multi Component Mesor

***Summary***: Rhythmicity adjusted activity mean based on a a multi-component Cosinor model. Measured as PIR Activity [s].

***Measure Ids***

1. *multi_component_mesor*
2. *multi_component_mesor_normalized*

***Type***

1. *Rhythmicity*

**2.1.5.1 Background and Hypothesis** The activity mesor $M$ is a rhythmicity adjusted activity mean, obtainable by means of multi-component cosinor curve fitting. Compared to the single component variant in 2.1.4, a multi-component model can better capture more complex activity cycles. . As with the single-component analog, here higher values refer to higher physical activity levels.

**2.1.5.2 Description** As with the single component variant, the mesor $M$ in equation (2) is directly estimated by fitting the respective model. The procedure to arrive at the mesor is the same as for the other cosinor derived parameters, see algorithm 2 for details.

###### 2.1.6 Cosinor Single Component Amplitude

***Summary***: Characterizes the strength or peakedness of a fitted single-component Cosinor regression model. Measured as PIR Activity [s].

***Measure Ids***

1. *one_component_amplitude*
2. *one_component_amplitude_normalized*

***Type***

1. *Rhythmicity*

**2.1.6.1 Background and Hypothesis** The activity amplitude is another parameter that can be estimated using cosinor analysis. It characterises the strength of the predictable activity rhythm. As with other cosinor parameters, we estimate it based on in-home PIR activity with the same justification elucidated in 2.1.2. Higher values here should be interpreted as a person having higher, more-pronounced activity rhythms.

**2.1.6.2 Description** In the single component case, the amplitude $A = \sqrt{A_1^2 + A_2^2}$ can be analytically calculated from the fit cosinor model, described in equation (1) [Moškon, 2020]. As described in [Li et al., 2020], to alleviate the impact of activity level differences between people, we calculate in addition to the regular amplitude a normalized one. In the normalized case, the amplitude is divided by the sample standard deviation of the person's activity data. The whole procedure is described in algorithm 2.

###### 2.1.7 Cosinor Multi Component Amplitude

***Summary***: A measure of activity cycle strength based on a multi-component Cosinor regression model. Measured as PIR Activity [s].

***Measure Ids***

1. *multi_component_amplitude*
2. *multi_component_amplitude_normalized*

***Type***

1. *Rhythmicity*

**2.1.7.1 Background and Hypothesis** See 2.1.2 for the basics. Compared to the single-component variant, the multi-component one is able to fit more complex activity curves and may thus lead to a more realistic estimate of the amplitude. Higher values here should be interpreted as a person having higher, more-pronounced activity rhythms.

**2.1.7.2 Description** As with the multi-component acrophase 2.1.3, the amplitude $A$ cannot be analytically calculated from the model (equation (2)), but must be estimated from the fitted curve. The whole procedure is described in algorithm 2.

###### 2.1.8 Cosinor Number of Components

***Summary***: A potential measure of activity cycle complexity. Measured as number of model components.

***Measure Ids***

1. *multi_component_n*

***Type***

1. *Rhythmicity*

**2.1.8.1 Background and Hypothesis** See 2.1.2 for the basics. The number of components refers to the number of components used for the multi-component cosionor model. We hypothesize that this may be to some degree representative of the complexity of the underlying activity cycles. As such, higher number of components may indicate more complex behavior patterns.

**2.1.8.2 Description** As described in 2.1.3, the optimal number of components $N$ in Equation (2), is found using the extra sum-of-squares F-test to compare competing models, taking into account the increased degrees of freedom when adding more components [Moškon, 2020]. We restricted the possible numbers of components to be between 1 and 6 components.

##### 2.1.9 Raw Fridge Usage

***Summary***: A measure of fridge usage frequency. Measured as number of fridge usages per day.

***Measure Ids***

1. *fridge_usage*

***Type***

1. *Activities of Daily Living*

**2.1.9.1 Background and Hypothesis** This is a measure describing the number of times a person's fridge was opened throughout a predefined time interval. We hypothesize that it may give insights into a person's eating and potentially drinking behavior. For instance, it is likely that an older adult that uses the fridge more regularly is more independent and potentially less frail. This may also be indicative of certain conditions leading to reduced appetite.

**2.1.9.2 Description** This is as simple as it sounds. We simply count the number of fridge opening events.

##### 2.1.10 Fridge Usage Epochs

***Summary***: A measure of fridge usage frequency. Measured as number of fridge usages per day.

***Measure Ids***

1. *fridge_usage_5min_epochs*

***Type***

1. *Activities of Daily Living*

**2.1.10.1 Background and Hypothesis** This is a simple measure similar to 2.1.9, describing the number of times a person's fridge was opened throughout a predefined time interval. However, as opposed to the raw version 2.1.9, we here calculate the opening based on 5 minute epochs, which may make the measure more generalizable. The rational for using fridge usage as a digital health measure is the same as in 2.1.9.

**2.1.10.2 Description** Here we first partition the fridge door openings into a set of non-overlapping 5-minute epochs containing the number of fridge openings inside a given epoch. The usage frequency is then defined as the cardinality of the subset of epochs with openings, defined by indicator function $I_{open}(x) := \begin{cases} 1 & \text{if } x > 0 \\ 0 & \text{otherwise} \end{cases}$, where $x$ is a scalar referring to the number of fridge openings.

##### 2.1.11 Daily Outing Number

***Summary***: A measure of outing frequency, thus how often a person leaves their home. Measured as number of outings per day.

***Measure Ids***

1. *outing_count*

***Type***

1. *Social Activities*
2. *Physical Activity*

**2.1.11.1 Background and Hypothesis** The outing number in a pre-defined time interval is a measure of how often a person leaves the apartment. Outing behavior may be related to physical activity levels and loneliness in older adults [Petersen et al., 2013]. Both, of which are linked to a variety of conditions including risk of mortality and cognitive decline.
We calculate outings based on heuristics as opposed to the more complex method proposed in [Petersen et al., 2013]. First of all, this does not require a ground-truth for training and should still be quite reliable given the door sensors are reliable - which we found to generally be the case with modern sensors (at least the ones we used).

**2.1.11.2 Description** As a first step, we create a list of potential outing intervals by segmenting time based on door-opening and closing pairs as for instance described in [Hu et al., 2017]. The rational here is that outings should occur between two entrance open/close event pairs. In a next step, data from all other sensors in a given apartment that clearly indicate the presence of a person (such as bed sensor or fridge sensor) are mapped to the previously extracted intervals. If any sensor event falls into a potential outing interval it is discarded. After this, PIR motion data is extracted and mapped to the remaining outing intervals. In a last step, all potential outing intervals with more than $4$ seconds of total PIR motion activity are removed - we found this threshold reasonable to deal with spurious PIR sensor activations that can occur from time to time. As a post-processing step, we discarded all outings values of days where practically ($\leq 5$ seconds) no PIR motion and no outing was detected at the same time (likely indicating sensor failure). For details see 3.

**Algorithm 3:** Calculate Daily Outings

```
function calculate_daily_outings (M, D, E);
Input: Matrix M containing PIR sensor firings with columns for time and duration of each activation, matrix D
  containing entrance door events with occurrence times and matrix E containing sensor events from other in-home
  sensors that mark presence (fridge sensor, bed sensor).
Output: Vector of daily outing number and total daily outing duration
MIN_PIR_ACTIVITY = 4;
Initialize list daily_outings;
outing_interval_candidates = calculate_openclose_intervals(D);
pir_motion_interval_tree = build_interval_tree(M);
other_inhome_sensor_interval_tree = build_interval_tree(E);
for candidate ∈ outing_interval_candidates do
    overlapping_pir_motion = pir_motion_interval_tree.query(candidate);
    overlapping_other_events = other_inhome_sensor_interval_tree.query(candidate);
    if sum(overlapping_pir_motion) > MIN_PIR_ACTIVITY || length(overlapping_other_events) > 0 then
        outing_interval_candidates.remove(candidate);
end
daily_outings = partition_into_days(outing_interval_candidates);
daily_motions = partition_into_days(M);
for i ∈ 0:length(daily_outings)-1 do
    motion_sum = sum(daily_motions[i]);
    outing_duration = sum(daily_outings[i].duration);
    if motion_sum >= 5 && outing_duration >= 5 then
        daily_outings.insert([length(daily_outings[i]), outing_duration])
end
return daily_outings;
```

##### 2.1.12 Daily Total Outing Duration

***Summary***: A measure of daily outing duration, thus how much time person spends outside the home. Measured as duration [s].

***Measure Ids***

1. *outing_duration*

***Type***

1. *Social Activities*
2. *Physical Activity*

**2.1.12.1 Background and Hypothesis** The background here is the same as in 2.1.11. The difference is that here we measure the total time spent outside the home.

**2.1.12.2 Description** Same as with 2.1.11 but summing over all individual outings of a given day. For details see 3.

##### 2.1.13 Daily Visit Score

***Summary***: A measure capturing the likelihood of a person receiving visits. Measured as a score, representing weighted visit seconds.

***Measure Ids***

1. *visit_score*

***Type***

1. *Social Activities*

**2.1.13.1 Background and Hypothesis** Home visits likely constitute an important part of social interaction in older adults. One would thus assume that higher visit scores would be indicative of more social interaction, less loneliness and potentially a reduced probability of having or developing associated conditions like late-life depression Schutz et al. [2021].

**2.1.13.2 Description** The daily visit score is calculated on the basis of a trained visit-detection algorithm introduced in Schutz et al. [2021]. Briefly, the PIR sensor events are segmented into non-overlapping segments based on door entrance opening-closing pairs, same as used for the outing detection 2.1.11. Subsequently a set of features is calculated based on the PIR sensor events and used in a self-training based domain adaptation algorithm to predict the probability of each segment being a visit or non-visit segment. The visit score for a given day is then given by the sum of individual segment scores $VS_i$ segments within a day. The segment scores $VS_i$ are defined as $VS_i = w_i \cdot |T_i|$, where $w_i$ refers to the prediction made by the trained visit-detection algorithm and $|T_i|$ represents the duration of a given segment. For more details see Schutz et al. [2021].

##### 2.1.14 Daily Number of Entrance Door Events

***Summary***: A measure of how often the entrance door has been opened and closed for a given day. Measured as number of entrance doors usages per day.

***Measure Ids***

1. *number_entrance_door_events*

***Type***

1. *Social Activities*
2. *Activities of Daily Living*

**2.1.14.1 Background and Hypothesis** Likely related to the outing number 2.1.11 but more basic. The number of entrance door openings should reflect both home visits and outings. It may thus capture some relation to loneliness but is likely less specific as the 2.1.13 or 2.1.11.

**2.1.14.2 Description** Simply the total number of all door events (opening and closing) of entrance doors that occurred throughout a day.

##### 2.1.15 Daily Time of Day Entrance Door

***Summary***: Time of day of first, middle and last occurrence of entrance door events for a given day. Measured as minutes since midnight [min].

***Measure Ids***

1. *entrance_door_tod_first*

2. *entrance_door_tod_middle*
3. *entrance_door_tod_last*

***Type***

1. *Activities of Daily Living*
2. *Social Activities*

**2.1.15.1 Background and Hypothesis** The time of day of specific events may measure certain behavioral tendencies and consistencies related to entrance door usage. The idea of using first, middle and last time of day occurrence of events has been used by Chen et al. [Chen et al., 2019].

**2.1.15.2 Description** Time of day of the first, middle and last entrance door event of a given day is simply calculated as the time of occurrence of a specific event converted to minutes since midnight (based on local time).

##### 2.1.16 Daily Time of Day Fridge Door

***Summary***: Time of day of first, middle and last occurrence of fridge door events for a given day. Measured as minutes since midnight [min].

***Measure Ids***

1. *fridge_door_tod_first*
2. *fridge_door_tod_middle*
3. *fridge_door_tod_last*

***Type***

1. *Activities of Daily Living*

**2.1.16.1 Background and Hypothesis** Same as in 2.1.15 but based on fridge door events. In this context, these measures may reflect upon eating behavior.

**2.1.16.2 Description** Time of day of the first, middle and last fridge door event of a given day is simply calculated as the time of occurrence of a specific event converted to minutes since midnight (based on local time).

##### 2.1.17 Daily Bathroom Usage Number

***Summary***: Contactless approximation of daily toilet usage. Measured as number of bathroom usage periods per day.

***Measure Ids***

1. *toilet_usage_number*

***Type***

1. *Activities of Daily Living*

**2.1.17.1 Background and Hypothesis** The idea here is to use PIR motion activity in the bathroom as an approximation for toilet usage. Toilet usage is not only related to specific conditions like urinary tract infections or congestive heart failure but could also be linked to cognitive function [Sugimoto et al., 2017]. As such, unusually high or low values may be indicative of underlying health issues.

**2.1.17.2 Description** While we cannot directly measure toilet usage with PIR sensors, we can approximate it using PIR activity in the bathroom. To do so, we calculate a set of non-overlapping 15-minute windows. Toilet usage can then be defined as the cardinality of the subset $\mathcal{T}_{active}$ of windows where PIR activity in the bathroom is greater than zero. This subset is defined by the indicator function $I_{\mathcal{T}_{active}}(\mathbf{x}) := \begin{cases} 1 & \text{if } sum(\mathbf{x}) > 0 \\ 0 & \text{otherwise} \end{cases}$, where $\mathbf{x}$ is a vector of PIR activity recordings in the bathroom for a given 15-minute window. To exclude days where a person was rarely at home or technical issues occurred, only days with a minimum total PIR activity of at least 300 seconds were included (similar to all other PIR based measures). To account for outings, intervals with outings were imputed by the rounded temporal average toilet usage occurrences when the person was at home. Imputation and filtering is the same as with other PIR based measures, see for instance algorithm 7.

###### 2.1.18 Bathroom Usage Time of Day

***Summary***: Contactless approximation of the time of day of first, middle and last toilet usage events. Measured as minutes since midnight [min].

***Measure Ids***

1. *toilet_usage_tod_first*
2. *toilet_usage_tod_middle*
3. *toilet_usage_tod_last*

***Type***

1. *Activities of Daily Living*

**2.1.18.1 Background and Hypothesis** Time of day of toilet usage events could measure certain aspects of toileting behavior that may be associated with cognitive processes or other urinary as well as self-care related conditions.

**2.1.18.2 Description** A PIR based approximation of toilet usage is calculated as described in 2.1.17. Using the subset $\mathcal{T}_{active}$ of windows with PIR activity in the bathroom, the time of occurrence of the first, middle and last event is calculated as the number of minutes after midnight (based on local time).

###### 2.1.19 Intradaily Variability

***Summary***: Contactless approximation of intradaily variability.

***Measure Ids***

1. *intradaily_variability*

***Type***

1. *Rhythmicity*

**2.1.19.1 Background and Hypothesis** intradaily variability (IV) is a measure of how fragmented daily activity patterns are [Gonçalves et al., 2014], where higher IV values indicate more fragmentation. It is a non-parametric measure often used with actigraphy data to analyse aspects of circadian rhythmicity in free-living conditions. Studies have for instance shown that IV may be indicative of sleep-wake cycle disturbances [Gonçalves et al., 2014]. In addition, there is research showing that higher IV values are linked to an increased risk of Alzheimer's dementia [Li et al., 2020, Gonçalves et al., 2014].

**2.1.19.2 Description** IV is a measure of how fragmented daily activity patterns are. We calculated it as described in [Li et al., 2020] but based on PIR sensor activity, instead of actigraphy counts

$$IV = \frac{n\sum_{i=2}^{n}(x_i - x_{i-1})^2}{(n-1)\sum_{i=1}^{n}(x_i - \bar{x})^2},$$

where $x_i$ is the sum of hourly PIR activity and $\bar{x}$ is the sample mean of $x_i$ over all hourly samples $n$. Activity for hours with outings were replaced with the temporal average of the same hour when the person was at home, multiplied by a factor of 1.4. Additionally, data from days with less than 300 seconds of total PIR activity, were removed. The last two steps are comparable to what is described in algorithm 6.

##### 2.1.20 Intradaily Stability

***Summary***: Contactless approximation of intradaily stability.

***Measure Ids***

1. *intradaily_stability*

***Type***

1. *Rhythmicity*

**2.1.20.1 Background and Hypothesis** intradaily stability (IS) is a measure of the robustness of daily (24h) activity rhythm [Li et al., 2020, Gonçalves et al., 2014]. It is a non-parametric measure often used with actigraphy data, where high IS values are indicative of good synchronization of the zeitgeber's 24 h cycle [Gonçalves et al., 2014]. It was shown that IS has a direct relationship with quality of life measures [Gonçalves et al., 2014]. Research also shows relationships with the Mini Mental State Examination (MMSE) [Gonçalves et al., 2014], a common questionnaire based measure for cognitive impairment, and that decreased IS is associated with an increased Alzheimer's dementia risk [Li et al., 2020].

**2.1.20.2 Description** IS is calculated based on a description in [Li et al., 2020] as follows

$$IS = \frac{n\sum_{h=1}^{24}(\bar{x}_h - \bar{x})^2}{24\sum_{i=1}^{n}(x_i - \bar{x})^2},$$

where $x_i$ is the sum of hourly PIR activity, $\bar{x}$ is the sample mean of $x_i$ over all hourly samples $n$ and $\bar{x}_h$ is the sample mean of activity across that specific hour of the day across all samples. Activity for hours with outings were replaced with the temporal average of the same hour when the person was at home, multiplied by a factor of 1.4. Additionally, data from days with less than 300 seconds of total PIR activity, were removed. The last two steps are comparable to what is described in algorithm 6.

##### 2.1.21 Spectral Entropy of Activity

***Summary***: Contactless measure of activity signal complexity. Measured as Shannon entropy [nats].

***Measure Ids***

1. *spectral_entropy*

***Type***

1. *Behavior Complexity*

**2.1.21.1 Background and Hypothesis** The spectral entropy (SE) of the activity signal can be viewed as a measure of complexity. Literature suggests that complexity (or inversely the lack of regularity) may be an adequate measure to quantify cognitive decline in older adults [Urwyler et al., 2017, Hayes et al., 2008, Chen et al., 2019]. Higher SE values indicate higher signal complexity.

**2.1.21.2 Description** In a first step, total PIR activity is binned (or resampled) into $N$ 1h periods $\mathbf{x}$. Subsequently, hours with outings are imputed by the temporal mean of the same hour when the person was at home, multiplied by a factor of 1.4, as with other PIR activity measures (see for instance 2.1.32). In a next step, the discrete Fourier transform (DFT) $\mathbf{X} = \mathcal{F}(\mathbf{x})$ is calculated. Then, the power spectral density of the transformed signal $\mathbf{X}$ is calculated and normalized $\hat{\mathbf{X}} = \dfrac{PSD(\mathbf{X})}{\sum PSD(\mathbf{X})}$, where the $PSD$ of the transform is calculated as

$$PSD(\mathbf{X}) = \frac{1}{N}|\mathbf{X}|^2. \tag{3}$$

Eventually, the SE is calculated as

$$SE = -\sum_{i=0}^{N-1} \hat{X}_i \log \hat{X}_i. \tag{4}$$

##### 2.1.22 LF-HF Power Ratio

***Summary***: Contactless measure of activity complexity. Measured as the ratio of low and high-frequency bands of the raw PIR sensor activity signal.

***Measure Ids***

1. *lfhf*

***Type***

1. *Behavior Complexity*

**2.1.22.1 Background and Hypothesis** The power spectral density ratio (power spectral density ratio (LFHF)) based on the PIR activity signal is similar to 2.1.21 in that we assume it may be a measure of behavioral complexity. The idea here is that people with regular behavior patterns will have higher relative power in lower frequency bands of the activity signal. On the contrary, non-periodic, erratic behaviour would more likely lead to higher relative power in the higher frequency bands. As a result, we would assume that larger LFHF values would indicate more regular behavior. The reasoning for attempting to quantify regularity in the signal is given in 2.1.21.

**2.1.22.2 Description** In a first step, total PIR activity is binned (or resampled) into $N$ 1h periods $\mathbf{x}$. Subsequently, hours with outings are imputed by the temporal mean of the same hour when the person was at home, multiplied by a factor of 1.4, as with other PIR activity measures (see for instance 2.1.32). In a next step, the DFT $\mathbf{X} = \mathcal{F}(\mathbf{x})$ is calculated. Then, the power spectral density $\bar{\mathbf{X}} = PSD(\mathbf{X})$ of the transformed signal $\mathbf{X}$ is calculated using equation (3).

For the calculation of the SE, only positive frequency bins $\{\frac{1}{2}N + 1, ..., N\}$ are considered with the low frequencies in the first half $\{\frac{1}{2}N, ..., \frac{3}{4}N\}$ and high frequencies in the second half $\{\frac{3}{4}N, ..., N\}$. Finally, LFHF is calculated as

$$LFHF = \frac{\sum_{i=N/2+1}^{N-N/4} \bar{X}_i}{\sum_{i=N-N/4+1}^{N} \bar{X}_i}.$$

where the frequency band is split in two evenly parts, for the low low and high frequencies, respectively. For simplicity, only positive frequencies are considered in the calculation.

##### 2.1.23 Wavelet Variance

***Summary***: Contactless approximation of activity variance across different timescales. Measured as normalized variance.

***Measure Ids***

1. *wavelet_variance_1.5h*
2. *wavelet_variance_3h*
3. *wavelet_variance_6h*
4. *wavelet_variance_12h*
5. *wavelet_variance_24h*

***Type***

1. *Behavior Complexity*

**2.1.23.1 Background and Hypothesis** Wavelet variance (wavelet variance (WV)) can be seen as a measure of variation in activity across different frequency scales. The basic assumption here is that the variance at a certain timescale may reflect upon the consistency of patterns at that scale. Higher WV values at a given timescale would thus rather indicate less predictable and more erratic behavior, as such WV tries to capture a similar relation as SE (2.1.21) or LFHF (2.1.22). Using wavelet variance in this context was introduced by Hayes et al. [Hayes et al., 2008], where they demonstrated that PIR based WV was higher in a mild cognitive impairment (MCI) group, compared to the healthy control group. Higher wavelet variance in a certain frequency band could be interpreted as more complex or variable behavior and less ordered behavior across the respective frequencies.

**2.1.23.2 Description** We calculate WV based on the description given in [Hayes et al., 2008] extended with our own pre-processing steps. First, we resample the activity signal into 11.25 minute bins. Subsequently we impute bins where outings occurred with the average activity of the same bin time of day without outing multiplied by a factor of 1.4 to account for higher activity outside the home (similar to how other PIR activity signals were handled, see for instance 2.1.32. Eventually we apply a energy normalized stationary wavelet transform (SWT) [Holschneider et al., 1990] to the signal and extract the detail coefficients at the following timescales: 45min - 1.5h, 1.5h - 3h, 3h - 6h, 6h - 12h, 12h - 24h. We should note that in the original description, it was not exactly specified whether the detail coefficients were calculated using regular discrete wavelet transform or stationary wavelet transform (or some related variant thereof [Fowler, 2005]). However, to get a meaningful estimate of wavelet variance the decimation of coefficients at each filtering step would lead to very short sequences, where edge effects are dominant, so we simply assume it was based on a non-decimated variant like SWT. For the SWT we use a coif5 wavelet function, as in [Hayes et al., 2008]. Based on the normalized detail coefficients we then calculate the variance of each coefficient sequence to obtain the final variance estimates. For the SWT calculations we used the PyWavelets implementation [Lee et al., 2019].

##### 2.1.24 Normalized Activity Bandpowers

***Summary***: Contactless approximation of activity signal bandpower in specific frequency bands. Measured as power spectral density $V^2/Hz$ and normalized power spectral density.

***Measure Ids***

1. *norm_amplitude_3e-06*
2. *norm_amplitude_1e-05*

...

20. *norm_amplitude_0.000129*

***Type***

1. *Behavior Complexity*
2. *Rhythmicity*

**2.1.24.1 Background and Hypothesis** The normalized PIR-activity bandpowers (PIR-BP) represent relative power in specific, evenly spaced frequency bands. Here the hypothesis is broad, as specific frequency bands may represent a variety of behaviors that may or may not be related to cognitive decline or a variety of other health indicators and outcomes.

**2.1.24.2 Description** We start out similarly as in 2.1.22. In a first step, total PIR activity is binned (or resampled) into $N$ 1h periods $\mathbf{x}$. Subsequently, hours with outings are imputed by the temporal mean of the same hour when the person was at home, multiplied by a factor of 1.4, as with other PIR activity measures (see for instance 2.1.32). In a next step, the DFT $\mathbf{X} = \mathcal{F}(\mathbf{x})$ is calculated. Then, the power spectral density $\bar{\mathbf{X}} = PSD(\mathbf{X})$ of the transformed signal $\mathbf{X}$ is calculated using equation (3). Now, the positive frequencies $N/2 : N-1$ are split into 20 evenly sized frequency bins $X_k^b \forall k \in [1, 20]$, ranging from 0 Hz to 0.00028 Hz (approximately 2h frequency). Each bin is then normalized by the total power across all bins, thus $\frac{X_k^b}{\sum_{i=0}^{20} X_i^b}$. Note that with this approach we discard shorter activity cycles, however, we think that those would hardly be comparable across subjects based on the already difficult to compare PIR signal and thus, at least for inter-subject comparison, would be less useful. Furthermore it should be mentioned that while we did not see strongly different values, the power spectral density may also be calculated using Welch's [Welch, 1967] method and made more comparable to other approaches by numerically integrating between power spectral density (PSD) points. However, as we did not see any larger differences between the more complicated and the presented simple approach, we use the simpler variant described above as that should be more intuitive to understand and potentially replicate.

##### 2.1.25 Fridge Usage Entropy

***Summary***: Shannon entropy measure of fridge usage, indicating uniformity of usage across the day. Measured as Shannon entropy [nats].

***Measure Ids***

1. *entropy_fridge*

***Type***

1. *Behavior Complexity*

###### 2.1.25.1 Background and Hypothesis

Fridge usage entropy (FE) tries to capture how even fridge usage is distributed across the day. We hypothesize that especially people with cognitive problems, like those with Alzheimer's dementia, would tend to use the fridge more evenly throughout the day leading to less clustered usage around specific times like local lunch or dinner times. Higher FE values indicate more uniformly distributed usage.

###### 2.1.25.2 Description

We calculate this measure by first counting fridge events for each hour of the day, giving us a vector $\mathbf{c} = [n_0, n_1, ..., n_{22}, n_{23}]$ where each component $n_h$ represents the count of fridge events for a given hour of the day $h$. Next we simply normalize the hourly counts to sum to 1, thus $\hat{\mathbf{c}} = \frac{\mathbf{c}}{\sum_{i=0}^{23} c_i}$. Lastly, FE is calculated based on classic Shannon entropy formulation

$$FE = -\sum_{i=0}^{23} \hat{c}_i \log \hat{c}_i$$

Here we exclude days that have less than 300 seconds of total PIR activation as with other estimates to exclude days with sensor issues or unusually long outings.

##### 2.1.26 Entrance Door Entropy

***Summary***: Shannon entropy measure of entrance door usage, indicating uniformity of usage across the day. Measured as Shannon entropy [nats].

***Measure Ids***

1. *entropy_entrance*

***Type***

1. *Behavior Complexity*

###### 2.1.26.1 Background and Hypothesis

Same as in 2.1.25 but based on entrance door usage.

###### 2.1.26.2 Description

Same as in 2.1.25 but based on entrance door events (openings/closings).

##### 2.1.27 PIR Activity Entropy

***Summary***: Shannon entropy measure of hourly PIR activity, indicating uniformity of activity across the day. Measured as Shannon entropy [nats].

***Measure Ids***

1. *total_entropy*

***Type***

1. *Behavior Complexity*

###### 2.1.27.1 Background and Hypothesis

Same as in 2.1.25 and 2.1.26 but on the basis of hourly PIR activity bins.

###### 2.1.27.2 Description

Overall the calculation is the same as in 2.1.25, with the exception of the raw PIR activity signal pre-processing. Here we first calculate hourly resampled activity bins and impute outings as with other PIR activity estimates (see 2.1.32). Based on those a vector containing the average activity levels per-hour of the day are calculated. After that the steps described in 2.1.25 are carried out, namely, the activity vector is normalized and Shannon entropy is calculated.

##### 2.1.28 Eigenbehavior Reconstruction Error

***Summary***: An eigendecomposition based measure of behavioral complexity. Measured as normalized error between input and reconstruction.

***Measure Ids***

1. *reconstruction_error*

***Type***

1. *Behavior Complexity*

**2.1.28.1 Background and Hypothesis** The eigenbehavior (or PCA) based reconstruction error (ERE) is attempting to calculate behavioral complexity of subjects based on PIR sensor derived location data. As with other measures of behavioral complexity like (2.1.21 or 2.1.22) the hypothesis is that cognitive decline may be measurable be exhibited behavioral complexity, thus how chaotic or irregular a person behaves in their daily lives, as for instance shown in [Hayes et al., 2008, Urwyler et al., 2017]. We introduced this measure in REF TO ANGELA and showed that higher ERE values are associated lower Montreal Cognitive Assessment (MoCA) scores. In general we would assume that ERE values indicate less structured behavior.

**2.1.28.2 Description** The approach is described in detail in REF TO ANGELA, here we give a brief overview. First a location matrix based on PIR sensor measurements $\mathbf{L} \in \mathbb{R}^{d \times 24|\mathcal{R}|}$ is calculated, where $d$ represents the number of days worth of location data and $\mathcal{R} \in \{kitchen, livingroom, toilet, bedroom, entrance, outside\}$ refers to the set of included locations. The row entries in $\mathbf{L}$ correspond to concatenated location vectors for each room, where for each hour of the day the percentage of room occupancy is encoded. Subsequently the centered location matrix $\hat{\mathbf{L}}$ is calculated from $\mathbf{L}$, which is then used to calculate the covariance matrix $\mathbf{\Sigma} = \hat{\mathbf{L}}(\hat{\mathbf{L}})^T$. Next, $\mathbf{\Sigma}$ is decomposed into a set of Eigenvalues and Eigenvectors using eigendecomposition. Based on the 7 Eigenvectors corresponding to the seven largest Eigenvalues, matrix $\bar{\mathbf{L}}$ is reconstructed. Eventually, the ERE is calculated as the mean sum of absolute differences between $\hat{\mathbf{L}}$ and $\bar{\mathbf{L}}$

$$ERE = \frac{1}{24d|\mathcal{R}|} \sum_{i=1}^{d} \sum_{j=1}^{24|\mathcal{R}|} \mathbf{D}_{i,j}, \; \mathbf{D} = |\hat{\mathbf{L}} - \bar{\mathbf{L}}|.$$

##### 2.1.29 Activity Island Number

***Summary:*** A contactless approximation of the number of physical activity bouts. Measured as number of activity islands per day

***Names:***

1. *activity_island_count*

***Type***

1. *Physical Activity*

**2.1.29.1 Background and Hypothesis** Islands of a measure can be thought of as continuous bursts or bouts of a certain activity and have been used as digital measures of health in [Chen et al., 2019]. In the context of physical activity, activity islands are conceptually similar to activity bouts as commonly extracted from accelerometer data in physical activity research (see for instance [Shiroma et al., 2019]). While PIR based activity islands are certainly not as accurate as accelerometer based ones, they likely still give some estimate on continuous physical activity occurrences. We assume that, compared to individual short PIR sensor activations, activity islands are more probable to capture moderate to vigorous physical activity levels and may thus give an different perspective compared to raw PIR based activity levels (2.1.32).
Physical activity in general, but especially higher levels thereof, is associated with a wider range of ageing relevant adverse health outcomes, including Alzheimer's disease, late-life depression, increased fall risk and all-cause mortality - to name a few.

**2.1.29.2 Description** To calculate activity islands, we first low-pass filter the raw PIR activity signal with a simple moving average filter and a window size of 2.5 seconds. Subsequently, all continuous activity stretches or "islands" are extracted. This means intervals where the filtered activity is consistently greater than 0 and the duration is at least 10 minutes. A more detailed description is given in algorithm 4.

**Algorithm 4:** Calculate PIR motion based Activity Islands

```
function calculate_pir_activity_islands (M, D, E);
Input: Matrix M containing PIR sensor firings with columns for time and duration of each activation, matrix D
  containing entrance door events with occurrence times and matrix E containing sensor events from other in-home
  sensors that mark presence (fridge sensor, bed sensor).
Output: Vector of daily activity island counts
Initialize list islands;
MIN_ACTIVITY = 300;
WINDOW_LENGTH_SECONDS = 150;
MIN_DURATION_SECONDS = 10 * 60;
filtered_activity = apply_moving_average(M, WINDOW_LENGTH_SECONDS);
activity_islands = extract_islands(filtered_activity, MIN_DURATION_SECONDS);
activity_days = partition_into_days(M);
islands_days = partition_into_days(activity_islands);
outings_days = calculate_outings(day, D, E);
for i ∈ 1:length(activity_days[i]) do
    motion = calculate_motion_sum(day);
    if motion > MIN_ACTIVITY then
        n_islands = count(islands_days[i]);
        median_island_duration = median(islands_days[i].duration);
        outing_estimate = calculate_temporal_count_medians(islands_days, outings_days);
        n_islands += outing_estimate;
        islands.insert([n_islands, median_island_duration]);
end
return islands;
```

##### 2.1.30 Activity Island Duration

***Summary***: A contactless approximation of the median length of physical activity bouts. Measured as duration [s].

***Measure Ids***

1. *activity_island_duration*

***Type***

1. *Physical Activity*

**2.1.30.1 Background and Hypothesis** This is the same as 2.1.29 but considering the median island duration instead of the count. We would assume that longer median activity bouts are indicative of higher overall fitness levels and comes with similar but potentially stronger associations with a multitude of health indicators and outcomes. However, literature is inconclusive on the importance of bout duration, with more recent work showing that it may not be as important [Jakicic et al., 2019].

**2.1.30.2 Description** Activity island duration is calculated with the same procedure as activity island number 2.1.29. Instead of counting the number of islands, we calculate the median duration of activity islands over a daily period.

##### 2.1.31 Activity Counts

***Summary***: A contactless approximation of the number of physical activity epochs. Measured as number of activity epochs per day

***Measure Ids***

1. *activity_counts*

***Type***

1. *Physical Activity*

**2.1.31.1 Background and Hypothesis** Another PIR motion based measure of in-home physical activity. The rational and hypothesis here is the same as in 2.1.32. The difference is in the way activity is calculated. Here, activity is based on the number of 5 minute epochs that have at least one sensor activation. It is thus less precise but potentially more comparable between apartments - without calibration.

**2.1.31.2 Description** PIR based activity counts are calculated by first partitioning the raw activity signals into a set of non-overlapping 5-minute epochs. The count is then given by the cardinality of the $active$ subset of epochs defined by the indicator function $I_{active}(\mathbf{x}) := \begin{cases} 1 & \text{if } sum(\mathbf{x}) > 0 \\ 0 & \text{otherwise} \end{cases}$, where $\mathbf{x}$ is a vector containing the raw activities of a given 5 minute epoch. Similar to other PIR based activity estimates, we impute outing intervals with the respective temporal mean of the same time period without outing, multiplied by a factor of 1.4 to account for higher activity outside the home [Schütz et al., 2020]. Days with unusually low activity (< 300 seconds) were excluded.

**Algorithm 5:** Calculate PIR Activity Count

```
function calculate_activity_counts (M, D, E);
Input: Matrix M containing PIR sensor firings with columns for time and duration of each activation, matrix D
  containing entrance door events with occurrence times and matrix E containing sensor events from other in-home
  sensors that mark presence (fridge sensor, bed sensor).
Output: Vector of daily activity count estimates
Initialize list daily_activity_counts;
MIN_ACTIVITY = 300;
outings = calculate_outings(M, D, E);
five_min_epochs = partition_into_5min_epochs(M);
activity = dichotomize(five_min_epochs);
binary_activity_epochs = partition_into_days(activity); daily_activity = partition_into_days(M); daily_outings =
  partition_into_days(outings);
for i ∈ 0:length(days)-1 do
    motion = calculate_motion_sum(day);
    if motion > MIN_ACTIVITY then
        outing_estimate = calculate_temporal_activity_means(binary_activity_epochs, daily_outings[i], outings);
        activity_count = sum(binary_activity_epochs[i]) + round(outing_estimate * 1.4);
        daily_activity_counts.insert(activity_count);
end
return daily_activity_counts;
```

##### 2.1.32 Raw Activity

***Summary***: A contactless approximation of physical activity. Measured as PIR activity duration [s].

***Measure Ids***

1. *raw_activity_sum_seconds*

***Type***

1. *Physical Activity*

**2.1.32.1 Background and Hypothesis** Passive infrared motion sensors might be used to approximately quantify in-home physical activity. One can thus interpret higher values as being associated with higher physical activity levels. In community-dwelling older adults, that spend a significant amount of time in their homes, this activity may be associated with overall physical activity, as we showed in a small cohort of older adults with the same apartment layout and sensor placement [Schütz et al., 2019]. We additionally showed that this value can be improved upon by imputing time spent outside the home with the temporal average of the same time interval across all monitored days, multiplied by a factor of approximately 1.4 [Schütz et al., 2020]. It should be noted, that the comparability of this measure between different apartments may be limited due to different apartment layouts and sensor placements, as well as a different number of sensors. To improve comparability, a calibration of PIR sensor systems may be employed if 7-14 days of accelerometer data is available [Schütz et al., 2020].

**2.1.32.2 Description** We calculated total raw activity by summing over all PIR sensor firing durations for a given day. Days with extremely little in-home activity (< 300 seconds) were excluded as they are likely the result of sensor issues or long outings. To reduce the effect of regular, short-term outings, we imputed activity during those outings with the temporal mean of these time intervals when the person was at home and multiplied this activity by a constant of 1.4 to account for the higher activity usually exerted during outings - for a justification see [Schütz et al., 2020]. A detailed description is given in Algorithm 6.

**Algorithm 6:** Calculate Raw PIR Sensor Activity

```
function raw_activity (M, D, E);
Input: Matrix M containing PIR sensor firings with columns for time and duration of each activation, matrix D
  containing entrance door events with occurrence times and matrix E containing sensor events from other in-home
  sensors that mark presence (fridge sensor, bed sensor).
Output: Vector of daily activity estimates
Initialize list daily_activities;
MIN_ACTIVITY = 300;
days = partition_into_days(M);
foreach day ∈ days do
    motion = calculate_motion_sum(day);
    if motion > MIN_ACTIVITY then
        outings = calculate_outings(day, D, E);
        outing_estimate = calculate_temporal_activity_means(days, outings, D);
        motion += outing_estimate;
        daily_activities.insert(motion);
end
return daily_activities;
```

##### 2.1.33 Daily Room-Transition Counts

***Summary***: A robust contactless approximation of physical activity. Measured as the number of room-transitions per day.

***Measure Ids***

1. *transition_count*

***Type***

1. *Physical Activity*

**2.1.33.1 Background and Hypothesis** This measure describes the number of times a person transitions between PIR sensor equipped rooms of an apartment. It can be seen as another, potentially more robust, way to quantify in-home physical activity [Schütz et al., 2019, Campbell et al., 2011]. In general it is plausible to assume that room-transitions of a certain type can be more comparable between subjects (compared to raw PIR based activity, without calibration), as it is less dependent on sensor positioning and apartment layout [Schütz et al., 2019]. We found this parameter to be associated with a variety of ageing relevant health-indicators and outcomes, at least in people sharing the same apartment layout [Schütz et al., 2019]. Work from Campbell et al. showed that room-transitions may be sensitive to health-status changes [Campbell et al., 2011].

**2.1.33.2 Description** The number of room-transitions is based on any transition (direct or indirect) between rooms that meets certain transition-time conditions: > 10 seconds and < 20 seconds. Formally, the number of transitions is the cardinality of the subset $\mathcal{R}_t$ of room-transition durations defined by the indicator function

$I_{\mathcal{R}_t}(x) := \begin{cases} 1 & \text{if } x > 10 \wedge x < 20 \\ 0 & \text{otherwise} \end{cases}$, where $x$ refers to a transition between any pair of rooms.

A transition duration is defined by $\Delta t$ between the first and last PIR firing times of two sensors in distinct rooms. To account for outings, we impute outing intervals by the expected count over the same interval when the person was at home.

While the duration constraint may seem a bit arbitrary, we found it to lead to more stable results. This is probably a result of excluding very short and potentially spurious transitions that are very much dependent on a specific room layout where a sensor may be triggered from another room. In addition, removing longer transitions makes sense as they do not represent a continuous motion but are the result of interwoven activities of daily living. For details see algorithm 7.

**Algorithm 7:** Calculate Room-Transitions

```
function calculate_room_transitions (M, D, E);
Input: Matrix M containing PIR sensor firings with columns for time and duration of each activation, matrix D
  containing entrance door events with occurrence times and matrix E containing sensor events from other in-home
  sensors that mark presence (fridge sensor, bed sensor). Assume all inputs are sorted by their time of occurrence.
Output: Vector of room-transition counts and average durations.
MIN_ACTIVITY = 300;
Initialize list daily_room_transitions;
Initialize list room_transitions;
for i ∈ 0:length(M) - 1 do
    dd t_activation_from = get_timestamp(M[i,:])
    room_from = get_room(M[i,:]);
    for j ∈ i:length(M) - 1 do
        room_to = get_room(M[j,:]);
        if room_from == room_to then
            continue;
        t_activation_to = get_timestamp(M[j,:]);
        Δt = t_activation_to - t_activation_from;
        if Δt <= 10 then
            continue;
        if Δt > 10 && Δt < 20 then
            room_transitions.insert(Δt);
        if Δt >= 20 then
            break;
    end
end
daily_activities = partition_into_days(M);
daily_outings = partition_into_days(calculate_outings(D, M, E));
daily_transitions = partition_into_days(room_transitions);
for i ∈ length(daily_activities) - 1 do
    motion = calculate_motion_sum(daily_activities[i]);
    if motion > MIN_ACTIVITY then
        outing_estimate = calculate_temporal_means(daily_outings[i], room_transitions, daily_outings);
        transition_count = count(daily_transitions[i]) + round(outing_estimate);
        transition_duration_mean = mean(daily_transitions[i]);
        daily_room_transitions.insert([transition_count, transition_duration_mean]);
end
return daily_room_transitions;
```

###### 2.1.34 Daily Mean Room-Transition Duration

***Summary***: A contactless approximation of gait-speed. Measured as duration [s].

***Measure Ids***

1. *mean_transition_duration*

***Type***

1. *Physical Activity*

**2.1.34.1 Background and Hypothesis** Similar to 2.1.33, however, here the duration between room-transitions is of interest. We assume that transition-duration could be used as an approximation of gait-speed [Kyrdalen et al., 2019, Piau et al., 2020] an important parameter related to fall risk but also Alzheimer's disease [Hayes et al., 2008, Buracchio et al., 2010]. However, without calibration, such as for instance shown in by Rana et al. [Rana et al., 2016], a PIR based approximation based on room-transitions may not generalize well beyond a single individual and could thus be of limited use for inter-subject comparisons.

**2.1.34.2 Description** Same as in 2.1.33, just that we are interested in the mean transition-duration $\bar{\mathcal{R}}_t$ of transition duration subset $\mathcal{R}_t$. For details see algorithm 7.

##### 2.1.35 Daily Mean Room-Transition Duration Morning

***Summary***: A contactless approximation of gait-speed in the morning. Measured as duration [s].

***Measure Ids***

1. *mean_morning_transition_duration*

***Type***

1. *Physical Activity*

**2.1.35.1 Background and Hypothesis** Same as 2.1.34 but with the restriction on morning hours (between 5:00 - 12:00). The rational here is that there is limited evidence [Hayes et al., 2008] that gait-speed may be different at different times of the day, which seems to have some association with MCI.

**2.1.35.2 Description** Same as 2.1.34 with the additional constraint that the transition event must fall in the time interval between 5:00 and 12:00.

##### 2.1.36 Daily Mean Room-Transition Duration Evening

***Summary***: A contactless approximation of gait-speed in the evening. Measured as duration [s].

***Measure Ids***

1. *mean_evening_transition_duration*

***Type***

1. *Physical Activity*

**2.1.36.1 Background and Hypothesis** Same idea as in 2.1.35. Here the transitions are restricted to evening hours (17:00 - 23:00).

**2.1.36.2 Description** Same as 2.1.34 with the additional constraint that the transition event must fall in the time interval between 5:00 and 12:00.

##### 2.1.37 Daily Mean Room-Transition Duration Ratio

***Summary***: A contactless approximation of the ratio of gait-speed in the morning vs evening. Measured as a ratio of morning and evening average room-transition durations.

***Measure Ids***

1. *morning_evening_transition_ratio*

***Type***

1. *Physical Activity*

**2.1.37.1 Background and Hypothesis** Same rational as in 2.1.35 and 2.1.36. However, here the ratio between morning and evening gait-speed of a given day is calculated.

**2.1.37.2 Description** The morning-evening gait-speed ratio is calculated by the ratio $\frac{\bar{\mathcal{R}}_t^{morning}}{\bar{\mathcal{R}}_t^{evening}}$, where the morning and evening refers to the transition-duration variants 2.1.35 and 2.1.36, respectively. For a detailed description of the transition-duration measure see 2.1.34 and 2.1.33.

##### 2.1.38 Daily Mean Room-Transition Duration Ratio Deviation

***Summary***: A contactless approximation of the deviation from an even ratio of gait-speed in the morning vs evening.

***Measure Ids***

1. *deviation_from_even_ratio*

***Type***

1. *Physical Activity*

**2.1.38.1 Background and Hypothesis** Same rational as 2.1.37. However, here only the deviation from an even gait-speed ratio is considered.

**2.1.38.2 Description** The morning-evening gait-speed ratio deviation is calculated by the absolute deviation from an even ratio of one $\left|1 - \frac{\bar{\mathcal{R}}_t^{morning}}{\bar{\mathcal{R}}_t^{evening}}\right|$, where the morning and evening refers to the average transition-duration variants 2.1.35 and 2.1.36, respectively. For a detailed description of the transition-duration measure see 2.1.34 and 2.1.33.

#### 2.2 Bed Mattress Sensor

##### 2.2.1 Nightly Heart Rate Summary Statistics

***Summary***: A set of robust in-bed heart rate summary statistic measures. Measured as heart beats per minute [bpm].

***Measure Ids***

1. *bed_hr_q10*
2. *bed_hr_q25*
3. *bed_hr_q50*
4. *bed_hr_q75*
5. *bed_hr_q90*
6. *bed_hr_iqr*
7. *bed_hr_kr3*
8. *bed_hr_sk3*

***Type***

1. *Sleep*

**2.2.1.1 Background and Hypothesis** Heart rate measures the heart's contraction frequency, which may serve as an indicator of autonomic nervous system activity and metabolic rate [Zhang and Zhang, 2009]. Similarly to daytime heart rate, it may to some degree related to physical fitness, stress, drugs as well certain diseases [Zhang and Zhang, 2009]. While in-bed heart rate often reflects resting heart rate, it is also influenced by sleep phases and may be lower than resting heart rate during deep sleep or higher during rapid eye movement (REM) sleep. One advantage of in-bed measurement is the comparability of measurements as environment and activity levels are somewhat similar and tend to be less modulated by external stimuli. In addition there is some evidence to suggest that increased night-time heart rate is associated with all-cause mortality and increased risk of cardiovascular disease events, while awake (or day-time) heart rate is not [Eguchi et al., 2009, Johansen et al., 2013].

**2.2.1.2 Description** We derive heart rate based on the manufacturer provided algorithms that extract beats from ballistocardiography signals and reports heart rate per minute every 4th second (0.25 Hz). The accuracy of the used algorithms was shown to be reasonable [Ranta et al., 2019]. The algorithms automatically mask invalid values as 0 or missing. For the statistics we omit those values as imputing them would likely lead to a bias as they are missing not at random but often as a result of motion, which we assume would itself likely influence heart rate and cannot realistically be estimated based on other variables in this case. We calculate the following quantiles: q10, q25, q50, q75, q90, representing the heart rate distribution throughout the night. In addition we calculate the inter quartile range (IQR) as a measure of dispersion. To quantify skewness and kurtosis we use the robust estimates SK3 and KR3, presented in [Kim and White, 2004].

##### 2.2.2 Nocturnal Heart Rate Dipping

***Summary***: An approximate measure of heart rate dipping during the night. Measured as quotient of the 75th and 25th heart rate [bpm] quantile throughout a night.

***Measure Ids***

1. *bed_hr_q75_q25*

***Type***

1. *Sleep*

**2.2.2.1 Background and Hypothesis** Absence of nocturnal decline in heart rate, also referred to as nocturnal dipping (ND) , was shown to be associated with all-cause mortality and risk of cardiovascular disease events [Eguchi et al., 2009].

**2.2.2.2 Description** We use the same heart rate signal, sampled at 0.25 Hz, described in 2.2.1. An approximation of ND is then calculated as the ratio between the first and third quartile

$$ND = \frac{Q_3}{Q_1}$$

##### 2.2.3 Nightly Heart Rate Average Power

***Summary***: Average power estimates of the nightly heart rate signal across different frequency bands. Measured as power spectral density [$V^2/Hz$] and normalized power spectral density.

***Measure Ids***

1. *bed_hr_total_power*
2. *bed_hr_bandpower_0.000000-0.002501_Hz*
3. *bed_hr_bandpower_0.002501-0.005003_Hz*

...

51. *bed_hr_bandpower_0.122568-0.124931_Hz*
52. *bed_hr_rel_bandpower_0.000000-0.002501_Hz*
53. *bed_hr_rel_bandpower_0.002501-0.005003_Hz*

...

101. *bed_hr_rel_bandpower_0.122568-0.124931_Hz*

***Type***

1. *Sleep*
2. *Sleep Complexity*

**2.2.3.1 Background and Hypothesis** Here we calculate average absolute and relative bandpower of the nightly heart rate signal. We hypothesize that certain heart rate patterns may be captured by specific frequency bands.

**2.2.3.2 Description** We use the same heart rate signal, sampled at 0.25 Hz, described in 2.2.1. Since we need continuous representations here, we interpolate missing values (although not ideal, as discussed in 2.2.1) with Akima interpolation [Akima, 1970]. The PSD of the heart rate signal for a given night is then calculated using Welch's method [Welch, 1967] with a "Hann" window of length 1799 (approximately 2 hours). As we already consider absolute heart rate values in 2.2.1, we subtract the signal mean prior to calculating the DFTs - no further detrending is performed. After the PSD is calculated, the PSD is dividied into 50 evenly spaced frequency bands from 0 to 0.125 Hz. For each band, as well as for the total spectrum, power is calculated as the numerical integral over the respective frequencies using the composite Simpson's rule. Additionally, relative bandpower for each band is calculated by dividing its power by the total power. Note that we consider only positive frequencies as we are dealing with real valued signals.

##### 2.2.4 Nightly Heart Rate Spectral Entropy

***Summary***: A complexity measure of the nightly heart rate signal. Measured as Shannon entropy [nats].

***Measure Ids***

1. *bed_hr_spectral_entropy*

***Type***

1. *Sleep Complexity*

**2.2.4.1 Background and Hypothesis** Complexity of nightly heart, or the lack thereof, could be an indicator of certain health conditions, although we do not have a specific hypothesis here, so this is really exploratory.

**2.2.4.2 Description** nightly heart rate spectral entropy (NHRSE) is calculated based on the same power spectral density estimates described in 2.2.3. We then calculate NHRSE by applying Shannon's entropy formulation to the additionally normalized PSD (normalized such that entries sum to one) using equation (4), where $\hat{X}_i$ refer to the normalized PSD entries.

##### 2.2.5 Nightly Respiration Rate Summary Statistics

***Summary***: A set of robust in-bed respiration rate summary statistic measures. Measured as respiration cycles per minute [rpm].

***Measure Ids***

1. *bed_rr_q10*
2. *bed_rr_q25*
3. *bed_rr_q50*
4. *bed_rr_q75*
5. *bed_rr_q90*
6. *bed_rr_iqr*
7. *bed_rr_kr3*
8. *bed_rr_sk3*

***Type***

1. *Sleep*

**2.2.5.1 Background and Hypothesis** Respiration rate at night can be indicative of certain medical conditions like congestive heart failure [Saner et al., 2021] or acute infections. It may also be affected by sleep-related breathing disorders like obstructive sleep apnea [Burman, 2017]. As with heart rate, respiration rate is influenced by different sleep stages and as such some variability throughout a night is expected [Chokroverty et al., 2010].

**2.2.5.2 Description** We derive respiration rate based on the manufacturer provided algorithms that extract breathing based on pressure differentials registered by a quasi-piezoelectric film placed under a person's mattress. As with nightly heart rate, the extracted respiration rate signal is reported once every 4th second (0.25 Hz). The accuracy of the used algorithms was shown to be reasonable [Ranta et al., 2019]. Further processing and calculation of the robust summary statistics are equivalent to what is described in 2.2.1.

##### 2.2.6 Nocturnal Respiration Rate Dipping

***Summary***: An approximate measure of respiration rate dipping during the night. Measured as quotient of the 75th and 25th respiration rate [rpm] quantile throughout a night.

***Measure Ids***

1. *bed_rr_q75_q25*

***Type***

1. *Sleep*

**2.2.6.1 Background and Hypothesis** Analogous to heart rate dipping we calculate this notion for respiration rate. However, we are not aware of a specific hypothesis or supporting literature.

**2.2.6.2 Description** We use the same respiration rate signal, sampled at 0.25 Hz, described in 2.2.5. An approximation of the decrease in respiration rate during the night is then calculated as the ratio between the first and third quartile.

$$NocturnalRespirationRateDipping = \frac{Q_3}{Q_1}$$

##### 2.2.7 Nightly Respiration Rate Average Power

***Summary***: Average power estimates of the nightly respiration rate signal across different frequency bands. Measured as power spectral density [$V^2/Hz$] and normalized power spectral density.

***Measure Ids***

1. *bed_rr_total_power*
2. *bed_rr_bandpower_0.000000-0.002501_Hz*
3. *bed_rr_bandpower_0.002501-0.005003_Hz*

...

51. *bed_rr_bandpower_0.122568-0.124931_Hz*
52. *bed_rr_rel_bandpower_0.000000-0.002501_Hz*
53. *bed_rr_rel_bandpower_0.002501-0.005003_Hz*

...

101. *bed_rr_rel_bandpower_0.122568-0.124931_Hz*

***Type***

1. *Sleep*
2. *Sleep Complexity*

**2.2.7.1 Background and Hypothesis** Here we calculate average absolute and relative bandpower of the nightly respiration rate signal. We hypothesize that certain respiration rate patterns may be captured by specific frequency bands.

**2.2.7.2 Description** The description here is exactly the same as with heart rate (see 2.2.3) but instead on the basis of the respiration rate signal, sampled at 0.25 Hz, as described in 2.2.5.

##### 2.2.8 Nightly Respiration Rate Spectral Entropy

***Summary***: A complexity measure of the nightly respiration rate signal. Measured as Shannon entropy [nats].

***Measure Ids***

1. *bed_rr_spectral_entropy*

***Type***

1. *Sleep Complexity*

**2.2.8.1 Background and Hypothesis** Complexity of the respiration rate signal may be related to certain medical conditions like obstructive breathing disorders [Burman, 2017].

**2.2.8.2 Description** nightly respiration rate spectral entropy (NRRSE) is calculated exactly the same way as its heart rate based pendant NHRSE (see 2.2.4, but based on the respiration rate signal instead of the heart rate signal.

##### 2.2.9 Nightly Bed Activity Summary Statistics

***Summary***: A set of robust in-bed activity summary statistic measures. Measured as unitless quantity proportional to in-bed actvity.

***Measure Ids***

1. *bed_act_q10*
2. *bed_act_q25*
3. *bed_act_q50*
4. *bed_act_q75*
5. *bed_act_q90*
6. *bed_act_iqr*
7. *bed_act_kr3*
8. *bed_act_sk3*

***Type***

1. *Sleep*

**2.2.9.1 Background and Hypothesis** In-bed activity simply refers to how much movement was observed. This is a unitless value that is a manufacturer provided estimate proportional to the the voltage produced by respective movements, as such, large movements will produce higher activity values. Activity or movements in bed related to sleep quality and may be indicative of specific movement related sleep disorders like restless leg syndrome. In addition, research indicates that increased movement at night is often related to health status changes and may be indicative of a wide variety of underlying conditions [Schütz et al., 2021]. It should be noted that bed activity is generally changing during healthy sleep based on the respective sleep stages, as such some variability is expected.

**2.2.9.2 Description** Similarly to the heart rate (2.2.1) and respiration rate (2.2.5) signal, we use a 0.25 Hz sampled activity signal, provided by the manufacturer. The exact signal processing this signal is subject to is not disclosed - if any. Bed exit periods are removed from the signal as they may introduce a bias towards zero activity values.

##### 2.2.10 Nightly Bed Activity Average Power

***Summary***: Average power estimates of the nightly activity signal across different frequency bands. Measured as power spectral density $[V^2/Hz]$ and normalized power spectral density.

***Measure Ids***

1. *bed_act_total_power*
2. *bed_act_bandpower_0.000000-0.002501_Hz*
3. *bed_act_bandpower_0.002501-0.005003_Hz*

...

51. *bed_act_bandpower_0.122568-0.124931_Hz*
52. *bed_act_rel_bandpower_0.000000-0.002501_Hz*

53. *bed_act_rel_bandpower_0.002501-0.005003_Hz*

...

101. *bed_act_rel_bandpower_0.122568-0.124931_Hz*

***Type***

1. *Sleep*
2. *Sleep Complexity*

**2.2.10.1 Background and Hypothesis** Here we calculate average absolute and relative bandpower of the nightly bed activity signal. We hypothesize that certain activity patterns may be captured by specific frequency bands (e.g. restless leg movements that occur at certain frequencies).

**2.2.10.2 Description** The description here is mostly the same as with heart rate (see 2.2.3) but on the basis of the bed activity signal, sampled at 0.25 Hz, as described in 2.2.9. In contrast to how heart rate bandpower is calculated (2.2.3), we here do not have missing values across the night but do cut out bed-exit periods.

##### 2.2.11 Nightly Bed Activity Spectral Entropy

***Summary***: A complexity measure of the nightly bed activity signal. Measured as Shannon entropy [nats].

***Measure Ids***

1. *bed_act_spectral_entropy*

***Type***

1. *Sleep Complexity*

**2.2.11.1 Background and Hypothesis** Complexity of the bed activity signal may be related to certain movement-related sleep disorders like restless leg syndrome or in general sleep disturbances.

**2.2.11.2 Description** The nightly bed activity spectral entropy (NBASE) is calculated similarly as its heart rate based pendant (see 2.2.4), but based on the bed activity signal instead of the heart rate signal and with removal of bed-exit periods.

##### 2.2.12 In-Bed Activity Island Count

***Summary***: Represents the number of continuous, in-bed, activity bouts per night. Measured as number of in-bed activity islands.

***Measure Ids***

1. *bed_number_activity_islands*

***Type***

1. *Sleep*

**2.2.12.1 Background and Hypothesis** This is based on the in-bed activity signal described in 2.2.9. However, instead of continuous activity we here calculate the number of in-bed activity bouts, which may be easier to compare across subjects and potentially even different devices. The idea of islands is the same one as described in 2.1.29 on the basis of PIR sensor registered activity.

**2.2.12.2 Description** In-Bed activity islands are computed by first deriving activity states, according to Algorithm 8 from the continuous bed activity signal (as introduced in 2.2.9). Based on this motion *on-off* states, continuous stretches of activity are extracted, which represent activity islands. Eventually, the number of islands per night is calculated.

**Algorithm 8:** Calculation of in-bed activity states.

```
function calculate_bed_activity_states (S);
Input: Matrix S containing continuous in-bed activity measurements and respective occurrence times.
Output: Vector containing activity states.
Initialize list bed_activity_states;
WINDOW_LENGTH_SECONDS = 300;
filtered_activity = apply_moving_average(S, WINDOW_LENGTH_SECONDS);
q75_activity = calculate_quantile(filtered_activity, 0.75);
for i ∈ 1:length(filtered_activity) do
    if filtered_activity[i] > q75_activity then
        bed_activity_states.insert(1);
    else
        bed_activity_states.insert(0);
    end
end
return bed_activity_states;
```

###### 2.2.13 In-Bed Activity Island Duration Statistics

***Summary***: A set of robust statistics on the duration of nightly in-bed activity islands. Measured as number of in-bed activity islands.

***Measure Ids***

1. *q10_bed_activity_island_duration*
2. *q25_bed_activity_island_duration*
3. *q50_bed_activity_island_duration*
4. *q75_bed_activity_island_duration*
5. *q90_bed_activity_island_duration*
6. *iqr_bed_activity_island_duration*

***Type***

1. *Sleep*

**2.2.13.1 Background and Hypothesis** The idea here is comparable to 2.2.12. However, instead of extracting the number of activity islands, we calculate summary statistics of their durations over a given night. We hypothesize that different duration profiles might be related to different sleep disturbances and differences in sleep architecture.

**2.2.13.2 Description** We calculate activity islands of a given night as described in 2.2.12. Instead of just counting them, however, we extract the duration of each island and calculate the 0.1, 0.25, 0.5, 0.75 and 0.9 quantiles as well as the interquartile range of the contained durations.

###### 2.2.14 In-Bed Arousal Counts

***Summary***: Reflects the number of transitions from an inactive to active state - while in bed. Measured as the number of times a person switches from inactive to active state in bed.

***Measure Ids***

1. *n_arousal_transitions*

***Type***

1. *Sleep*

**2.2.14.1 Background and Hypothesis** The number of arousals refers to the total number a person transitions from an inactive activity state to an active in-bed activity state over a night of sleep. We hypothesize that this could measure, for instance, sleep fragmentation and therefore be indicative of sleep quality.

**2.2.14.2 Description** To calculate the arousal count (AC), we first calculate in-bed activity states based on algorithm 8. Subsequently, we calculate the number a person changes from an inactive to an active state.

##### 2.2.15 In-Bed Arousal Probability

***Summary***: Refers to the probability of transitioning from an inactive to an active in-bed activity state. Measured as transition probability.

***Measure Ids***

1. *arousal_probability*

***Type***

1. *Sleep*

**2.2.15.1 Background and Hypothesis** This measure is based on 2.2.14. Instead of calculating an absolute number of arousals, we here calculate the probability of transitioning from an inactive to an active in-bed activity state. This may be more objective and thus better comparable across subjects.

**2.2.15.2 Description** To in-bed arousal probability is calculated by first extracting the number of transitions from inactive to active in-bed activity state AC, as described in 2.2.14. Subsequently, the arousal probability is estimated by dividing AC by the sum of AC and the number of inactive to inactive state-transitions (NC): $Pr(arousal) = \frac{AC}{AC+NC}$. This is calculated for each night.

##### 2.2.16 Percentage Active in Bed

***Summary***: Describes the percentage of a night spent in an active in-bed activity state. Measured as percentage.

***Measure Ids***

1. *percentage_active_in_bed*

***Type***

1. *Sleep*

**2.2.16.1 Background and Hypothesis** Related to 2.2.15, the percentage active in bed, represents the percentage a person was in an active in-bed state for a given night. Contrary to 2.2.15, here the temporal aspect is not considered, instead we quantify the overall percentage. This measure should be related to sleep quality we expect it to correlate with time spent awake in bed.

**2.2.16.2 Description** Here we first extract the in-bed activity states based on algorithm 8. Thereafter, we simply divide the total number of active states by the total number of in-bed activity states for a given night.

##### 2.2.17 Duration Total

***Summary***: Total duration of a nightly sleep episode. Measured as duration [s].

***Measure Ids***

1. *bed_duration*

***Type***

1. *Sleep*

**2.2.17.1 Background and Hypothesis** The total duration of a sleep episode refers to the total duration, in seconds, from the first to the last registered sleep entry. Note that this includes also shorter bed-exits, such as nightly toilet visits.

**2.2.17.2 Description** This measure is extracted by algorithms from the device manufacturer.

###### 2.2.18 Duration In-Bed

***Summary***: Total duration spent in bed per night. Measured as duration [s].

***Measure Ids***

1. *bed_duration_in_bed*

***Type***

1. *Sleep*

**2.2.18.1 Background and Hypothesis** The total duration spent in bed refers to the total amount of seconds spent in bed. This excludes bed-exits.

**2.2.18.2 Description** This measure is extracted by algorithms from the device manufacturer.

###### 2.2.19 Nightly Bed-exit Count

***Summary***: The number of bed-exits of a night.

***Measure Ids***

1. *bed_bedexit_count*

***Type***

1. *Sleep*

**2.2.19.1 Background and Hypothesis** Measures the total number of bed-exits of a single night record. This may be related to activities like nocturia. Especially with ageing this number is expected to increase but can also be related to underlying conditions like bladder infections or congestive heart failure [Saner et al., 2021, Bosch and Weiss, 2010]. In addition, nightly bed exits in older adults can be a risk factor for falls [Stewart et al., 1992, Awais et al., 2019].

**2.2.19.2 Description** This measure is extracted by algorithms from the device manufacturer.

###### 2.2.20 Number of Toss-and-Turns per Night

***Summary***: Quantifies the number of larger body movements throughout the night.

***Measure Ids***

1. *bed_tossnturn_count*

***Type***

1. *Sleep*

**2.2.20.1 Background and Hypothesis** This measure represents the number of larger body movement occurrences over a given night. Increased body movements may be related to restless and more fragmented sleep. We showed that this measure is oftentimes a highly relevant early sign for health deteriorations in older adults [Schütz et al., 2021].

**2.2.20.2 Description** This measure is extracted by algorithms from the device manufacturer.

###### 2.2.21 Average In-Bed Heart Rate

***Summary***: The average heart rate while in bed. Measured as number of heart beats per minute [bpm].

***Measure Ids***

1. *bed_avg_hr*

***Type***

1. *Sleep*

**2.2.21.1 Background and Hypothesis** This is mostly equal to 2.2.1 but based on calculations from the device manufacturer.

**2.2.21.2 Description** This measure is extracted by algorithms from the device manufacturer.

##### 2.2.22 Average In-Bed Respiration Rate

***Summary***: The average respiration rate while in bed. Measured as number of respiration cycles per minute [rpm].

***Measure Ids***

1. *bed_avg_rr*

***Type***

1. *Sleep*

**2.2.22.1 Background and Hypothesis** This is mostly equal to 2.2.5 but based on calculations from the device manufacturer.

**2.2.22.2 Description** This measure is extracted by algorithms from the device manufacturer.

##### 2.2.23 Average In-Bed Activity

***Summary***: The average activity while in bed.

***Measure Ids***

1. *bed_avg_act*

***Type***

1. *Sleep*

**2.2.23.1 Background and Hypothesis** This is mostly equal to 2.2.9 but based on calculations from the device manufacturer.

**2.2.23.2 Description** This measure is extracted by algorithms from the device manufacturer.

##### 2.2.24 Nightly Duration Asleep

***Summary***: The number of seconds spent asleep for a given night-segment. Measured as duration [s].

***Measure Ids***

1. *bed_duration_in_sleep*

***Type***

1. *Sleep*

**2.2.24.1 Background and Hypothesis** This measure represents the total time (in seconds) a person was asleep for a night. Adequate sleep time is important for maintaining both mental and physical well-being. Too short sleep is associated with a variety of negative health outcomes, including increased mortality, cardiovascular disease, hypertension and obesity [Itani et al., 2017]. Similar associations were found regarding long sleep durations [Jike et al., 2018]. Furthermore, there is evidence suggesting a relationship between sleep duration and dementia related neurodegeneration [Westwood et al., 2017].

**2.2.24.2 Description** This measure is extracted by algorithms from the device manufacturer.

###### 2.2.25 Nightly Duration Awake In-Bed

***Summary***: The time spent awake in-bed for a given night-segment. Measured as duration [s].

***Measure Ids***

1. *bed_duration_awake*

***Type***

1. *Sleep*

**2.2.25.1 Background and Hypothesis** This measure represents the total time (in seconds) a person was awake, while in bed, for a given night. Being awake in-bed for longer durations may be related to sleep disorders like insomnia, which in turn is associated with a wide variety of conditions, such as depression or chronic illnesses [Roth, 2007].

**2.2.25.2 Description** This measure is extracted by algorithms from the device manufacturer.

###### 2.2.26 Sleep Onset Delay

***Summary***: The time it takes to fall asleep. Measured as duration [s].

***Measure Ids***

1. *bed_duration_sleep_onset*

***Type***

1. *Sleep*

**2.2.26.1 Background and Hypothesis** This measure represents the total time (in seconds) it took for a person to fall asleep. Similar to 2.2.25 longer sleep onset delay is often related to insomnia, with the same implications as described in 2.2.25.

**2.2.26.2 Description** This measure is extracted by algorithms from the device manufacturer.

###### 2.2.27 Nightly Bedexit Duration

***Summary***: The time spent outside the bed during a sleep period. Measured as duration [s].

***Measure Ids***

1. *bed_bedexit_duration*

***Type***

1. *Sleep*

**2.2.27.1 Background and Hypothesis** This measure refers to the time spent outside the bed during a recorded sleep period.

**2.2.27.2 Description** This measure is extracted by algorithms from the device manufacturer.

###### 2.2.28 Nightly REM Sleep Duration

***Summary***: The total duration per night spent in rapid eye movement sleep. Measured as duration [s].

***Measure Ids***

1. *bed_duration_in_rem*

***Type***

1. *Sleep*

**2.2.28.1 Background and Hypothesis** This measure refers to the time (in seconds) spent in REM sleep. REM sleep is still not very well understood but is thought to be important for memory consolidation and learning [Peever and Fuller, 2017].

**2.2.28.2 Description** This measure is extracted by algorithms from the device manufacturer.

##### 2.2.29 Nightly Light Sleep Duration

***Summary***: The total duration per night spent in light sleep. Measured as duration [s].

***Measure Ids***

1. *bed_duration_in_light*

***Type***

1. *Sleep*

**2.2.29.1 Background and Hypothesis** This measure refers to the total time (in seconds) spent in the light sleep stage (N1 and N2 or stage 1 and stage 2) over the course of a night. Light sleep is usually encountered after being awake or shortly before awakening. Older adults tend to be more in this stage of sleep, which is part of the reason why they are more likely to wake up throughout the night [Cooke and Ancoli-Israel, 2006].

**2.2.29.2 Description** This measure is extracted by algorithms from the device manufacturer.

##### 2.2.30 Nightly Deep Sleep Duration

***Summary***: The total duration per night spent in deep sleep. Measured as duration [s].

***Measure Ids***

1. *bed_duration_in_deep*

***Type***

1. *Sleep*

**2.2.30.1 Background and Hypothesis** This measure refers to the amount of time (in seconds) a person spent in deep sleep (slow wave sleep, delta sleep, N3, or stage 3) throughout a given sleep record. With ageing the percentage of deep sleep tends to decrease [Cooke and Ancoli-Israel, 2006].

**2.2.30.2 Description** This measure is extracted by algorithms from the device manufacturer.

##### 2.2.31 Nightly Heart Rate Variability LF

***Summary***: Heart rate variability power in the low frequency component of the HRV spectral distribution. Measured as normalized power spectral density.

***Measure Ids***

1. *bed_hrv_lf*

***Type***

1. *Sleep*

**2.2.31.1 Background and Hypothesis** This measure summarizes the power in the low frequency band (0.04-0.15 Hz) of heart rate variability (HRV LF) throughout a given night. Heart rate variability is the variability in the time difference between consecutive heart beats. The measure is normalized such that low frequency power and high frequency power sum to 100. During resting conditions, low frequency (0.04–0.15 Hz) (LF) power primarily reflects on baroreceptor activity [Shaffer and Ginsberg, 2017].

**2.2.31.2 Description** This measure is extracted by algorithms from the device manufacturer.

##### 2.2.32 Nightly Heart Rate Variability HF

***Summary***: Heart rate variability power in the high frequency component of the HRV spectral distribution. Measured as normalized power spectral density.

***Measure Ids***

1. *bed_hrv_hf*

***Type***

1. *Sleep*

**2.2.32.1 Background and Hypothesis** This measure summarizes the power in the high frequency band (0.15-0.40 Hz) of heart rate variability (HRV HF) throughout a given night. Heart rate variability is the variability in the time difference between consecutive heart beats. The measure is normalized such that low frequency power and high frequency power sum to 100. HRV HF tends to be higher at night than during the day. It is thought to reflect parasympathetic activity and is influenced by respiratory cycles [Shaffer and Ginsberg, 2017]. Furthermore, HRV HF is often highly correlated with the commonly used HRV measures, pNN50 and RMSSD, and was found to be correlated with stress, panic, anxiety or worry [Shaffer and Ginsberg, 2017].

**2.2.32.2 Description** This measure is extracted by algorithms from the device manufacturer.

##### 2.2.33 Number of Awakenings per Night

***Summary***: The number of awakenings during a sleep period.

***Measure Ids***

1. *bed_awakenings*

***Type***

1. *Sleep*

**2.2.33.1 Background and Hypothesis** Represents the number of times a person woke up throughout the night. This should be related to the quality of sleep and may be indicative of certain sleep disorders like obstructive sleep apnea.

**2.2.33.2 Description** This measure is extracted by algorithms from the device manufacturer.

##### 2.2.34 Percentage in Deep Sleep

***Summary***: The percentage spent in deep sleep over the night.

***Measure Ids***

1. *bed_percentage_deep*

***Type***

1. *Sleep*

**2.2.34.1 Background and Hypothesis** This measure represents the percentage of time spent in deep sleep (2.2.30) with respect to the overall sleep duration (2.2.18) $\frac{t_{deep\_sleep}}{t_{bed}}$.

**2.2.34.2 Description** This measure is calculated based on the respective measures extracted by algorithms from the device manufacturer.

###### 2.2.35 Percentage in Light Sleep

***Summary***: The percentage spent in light sleep over the night.

***Measure Ids***

1. *bed_percentage_light*

***Type***

1. *Sleep*

**2.2.35.1 Background and Hypothesis** This measure represents the percentage of time spent in light sleep (2.2.29) with respect to the overall sleep duration (2.2.18) $\frac{t_{light\_sleep}}{t_{bed}}$.

**2.2.35.2 Description** This measure is calculated based on the respective measures extracted by algorithms from the device manufacturer.

###### 2.2.36 Percentage in REM Sleep

***Summary***: The percentage spent in rem sleep over the night.

***Measure Ids***

1. *bed_percentage_rem*

***Type***

1. *Sleep*

**2.2.36.1 Background and Hypothesis** This measure represents the percentage of time spent in REM sleep (2.2.28) with respect to the overall sleep duration (2.2.18) $\frac{t_{rem\_sleep}}{t_{bed}}$.

**2.2.36.2 Description** This measure is calculated based on the respective measures extracted by algorithms from the device manufacturer.

###### 2.2.37 Percentage Awake

***Summary***: The percentage spent awake with respect to the sleep record.

***Measure Ids***

1. *bed_percentage_awake*

***Type***

1. *Sleep*

**2.2.37.1 Background and Hypothesis** This measure represents the percentage of time spent in awake (2.2.25) with respect to the overall sleep duration (2.2.18) $\frac{t_{awake}}{t_{bed}}$.

**2.2.37.2 Description** This measure is calculated based on the respective measures extracted by algorithms from the device manufacturer.

###### 2.2.38 Sleep Efficiency

***Summary***: The percentage of time spent asleep with respect to overall time spent in the bed.

***Measure Ids***

1. *bed_sleep_efficiency*

***Type***

1. *Sleep*

**2.2.38.1 Background and Hypothesis** This measure represents the percentage of time spent asleep (2.2.24) with respect to the duration in bed (2.2.18) $\frac{t_{asleep}}{t_{bed}}$. Sleep efficiency is a commonly used quality of sleep metric, where lower values indicate worse quality sleep. It has been shown that sleep efficiency decreases with ageing, however, this decrease may not be universal and older adults with higher sleep efficiency tend to have less health issues [Didikoglu et al., 2020].

**2.2.38.2 Description** This measure is calculated based on the respective measures extracted by algorithms from the device manufacturer.

###### 2.2.39 Bed-exit Duration Statistics

***Summary***: Median of value of bed-exit durations for a given night. Measured as duration [s].

***Measure Ids***

1. *bed_bedexit_duration_median*

***Type***

1. *Sleep*

**2.2.39.1 Background and Hypothesis** This is the same as 2.2.27 but reporting the median of individual exits (if multiple exits occurred) instead of the total bed-exit duration across a given sleep record.

**2.2.39.2 Description** This measure is calculated based on the respective measures extracted by algorithms from the device manufacturer.

###### 2.2.40 Nightly Nocturia Count

***Summary***: Number toilet events during a given sleep record.

***Measure Ids***

1. *bed_nocturia_count*

***Type***

1. *Sleep*

**2.2.40.1 Background and Hypothesis** This represents the number of nightly toilet visits that occurred throughout a sleep record. Nocturia is associated with increased fall risk [Stewart et al., 1992] but can also be an indicator for underlying conditions like congestive heart failure [Saner et al., 2021, Ohishi et al., 2021].

**2.2.40.2 Description** Here we used the manufacturer provided bed-exit times together with PIR sensor readings of a sensor in the bathroom. If a bed-exit coincided with a bathroom activation, the event is counted as nocturia occurrence.

###### 2.2.41 Average Nightly Heart Rate during Awake State

***Summary***: Average heart rate value when awake in bed. Measured as heart beats per minute [bpm].

***Measure Ids***

1. *bed_awake_sleep_hr*

***Type***

1. *Sleep*

**2.2.41.1 Background and Hypothesis** This represents the average in-bed heart rate of a person while being awake during a given sleep record.

**2.2.41.2 Description** We calculated this measure using the manufacturer provided sleep stage information together with the continuous heart rate signal introduced in 2.2.1.

###### 2.2.42 Average Nightly Respiration Rate during Awake State

***Summary***: Average respiration rate value when awake in bed. Measured as respiration cycles per minute [rpm].

***Measure Ids***

1. *bed_awake_sleep_rr*

***Type***

1. *Sleep*

**2.2.42.1 Background and Hypothesis** This represents the average in-bed respiration rate of a person while being awake during a given sleep record.

**2.2.42.2 Description** We calculated this measure using the manufacturer provided sleep stage information together with the continuous respiration rate signal introduced in 2.2.5.

###### 2.2.43 Average Nightly Activity during Awake State

***Summary***: Average in-bed activity when awake in bed.

***Measure Ids***

1. *bed_awake_sleep_act*

***Type***

1. *Sleep*

**2.2.43.1 Background and Hypothesis** This represents the average in-bed activity of a person while being awake during a given sleep record.

**2.2.43.2 Description** We calculated this measure using the manufacturer provided sleep stage information together with the continuous activity signal introduced in 2.2.9.

###### 2.2.44 Average Nightly Heart Rate during Deep Sleep

***Summary***: Average heart rate value while in deep sleep. Measured as the number of heart beats per minute [bpm].

***Measure Ids***

1. *bed_deep_sleep_hr*

***Type***

1. *Sleep*

**2.2.44.1 Background and Hypothesis** This represents the average in-bed heart rate of a person while being in deep sleep, during a given sleep record.

**2.2.44.2 Description** We calculated this measure using the manufacturer provided sleep stage information together with the continuous heart rate signal introduced in 2.2.1.

###### 2.2.45 Average Nightly Respiration Rate during Deep Sleep

***Summary***: Average respiration rate value while in deep sleep. Measured as the number of respiration cycles per minute [rpm].

***Measure Ids***

1. *bed_deep_sleep_rr*

***Type***

1. *Sleep*

**2.2.45.1 Background and Hypothesis** This represents the average in-bed respiration rate of a person while being in deep sleep, during a given sleep record.

**2.2.45.2 Description** We calculated this measure using the manufacturer provided sleep stage information together with the continuous respiration rate signal introduced in 2.2.5.

##### 2.2.46 Average Nightly Activity during Deep Sleep

***Summary***: Average in-bed activity value while in deep sleep.

***Measure Ids***

1. *bed_deep_sleep_act*

***Type***

1. *Sleep*

**2.2.46.1 Background and Hypothesis** This represents the average in-bed activity value of a person while being in deep sleep, during a given sleep record.

**2.2.46.2 Description** We calculated this measure using the manufacturer provided sleep stage information together with the continuous activity signal introduced in 2.2.9.

##### 2.2.47 Average Nightly Heart Rate during Light Sleep

***Summary***: Average heart rate value while in light sleep. Measured as the number of heart beats per minute [bpm].

***Measure Ids***

1. *bed_light_sleep_hr*

***Type***

1. *Sleep*

**2.2.47.1 Background and Hypothesis** This represents the average in-bed heart rate of a person while being in light sleep, during a given sleep record.

**2.2.47.2 Description** We calculated this measure using the manufacturer provided sleep stage information together with the continuous heart rate signal introduced in 2.2.1.

##### 2.2.48 Average Nightly Respiration Rate during Light Sleep

***Summary***: Average respiration rate value while in light sleep. Measured as the number of respiration cycles per minute [rpm].

***Measure Ids***

1. *bed_light_sleep_rr*

***Type***

1. *Sleep*

**2.2.48.1 Background and Hypothesis** This represents the average in-bed respiration rate of a person while being in light sleep, during a given sleep record.

**2.2.48.2 Description** We calculated this measure using the manufacturer provided sleep stage information together with the continuous respiration rate signal introduced in 2.2.5.

##### 2.2.49 Average Nightly Activity during Light Sleep

***Summary***: Average in-bed activity value while in light sleep.

***Measure Ids***

1. *bed_light_sleep_act*

***Type***

1. *Sleep*

**2.2.49.1 Background and Hypothesis** This represents the average in-bed activity value of a person while being in light sleep, during a given sleep record.

**2.2.49.2 Description** We calculated this measure using the manufacturer provided sleep stage information together with the continuous activity signal introduced in 2.2.9.

##### 2.2.50 Average Nightly Heart Rate during REM Sleep

***Summary***: Average heart rate value while in REM sleep. Measured as the number of heart beats per minute [bpm].

***Measure Ids***

1. *bed_rem_sleep_hr*

***Type***

1. *Sleep*

**2.2.50.1 Background and Hypothesis** This represents the average in-bed heart rate of a person while being in REM sleep, during a given sleep record.

**2.2.50.2 Description** We calculated this measure using the manufacturer provided sleep stage information together with the continuous heart rate signal introduced in 2.2.1.

##### 2.2.51 Average Nightly Respiration Rate during REM Sleep

***Summary***: Average respiration rate value while in REM sleep. Measured as the number of respiration cycles per minute [rpm].

***Measure Ids***

1. *bed_rem_sleep_rr*

***Type***

1. *Sleep*

**2.2.51.1 Background and Hypothesis** This represents the average in-bed respiration rate of a person while being in REM sleep, during a given sleep record.

**2.2.51.2 Description** We calculated this measure using the manufacturer provided sleep stage information together with the continuous respiration rate signal introduced in 2.2.5.

##### 2.2.52 Average Nightly Activity during REM Sleep

***Summary***: Average in-bed activity value while in REM sleep.

***Measure Ids***

1. *bed_rem_sleep_act*

***Type***

1. *Sleep*

**2.2.52.1 Background and Hypothesis** This represents the average in-bed activity value of a person while being in REM sleep, during a given sleep record.

**2.2.52.2 Description** We calculated this measure using the manufacturer provided sleep stage information together with the continuous activity signal introduced in 2.2.9.

##### 2.2.53 Sleep Reconstruction Error

***Summary***: Eigendecomposition based measure of sleep behavior complexity. Measured as normalized error between input and reconstruction.

***Measure Ids***

1. *reconstruction_error_1*
2. *reconstruction_error_2*

***Type***

1. *Sleep Complexity*

**2.2.53.1 Background and Hypothesis** This represents a sleep structure complexity measure over multiple days and is based on ERE as introduced in 2.1.28. The hypothesis being that more regular sleeping behavior, thus lower reconstruction errors, should be associated with better health outcomes.

**2.2.53.2 Description** The approach here is comparable to the very similar measure based on PIR locations 2.1.28. However, in this context matrix $\mathbf{L}$, represents different sleep stages and associated heart rate, respiration rate, as well as raw activity. $\mathbf{L} \in \mathbb{R}^{d \times 24|\mathcal{S}|}$ is calculated, where $d$ represents the number of days worth of sleep data and $\mathcal{S} \in \{outofbed, awake, lightsleep, deepsleep, remsleep, heartrate, respirationrate, activity\}$ refers to the respective sleep measurements. The row entries in $\mathbf{L}$ correspond to concatenated measurement vectors for each sleep stage, where for each half hour recording up to 12 hours, the percentage in the respective state is encoded. In case of heart rate, respiration rate and activity, the mean values are encoded. Subsequently the centered location matrix $\hat{\mathbf{L}}$ is calculated from $\mathbf{L}$, which is then used to calculate the covariance matrix $\mathbf{\Sigma} = \hat{\mathbf{L}}(\hat{\mathbf{L}})^T$. Next, $\mathbf{\Sigma}$ is decomposed into a set of Eigenvalues and Eigenvectors using eigendecomposition. Based on the two Eigenvectors corresponding to the two largest Eigenvalues, the matrix $\bar{\mathbf{L}}_1$ (based on the largest Eigenvalue) and $\bar{\mathbf{L}}_2$ (based on the largest two Eigenvalues) are reconstructed. Eventually, the sleep reconstruction errors (SRE) based on one or two Eigenvectors is calculated as the mean sum of absolute differences between $\hat{\mathbf{L}}$ and $\bar{\mathbf{L}}_{\{1,2\}}$

$$ERE_{\{1,2\}} = \frac{1}{24d|\mathcal{S}|} \sum_{i=1}^{d} \sum_{j=1}^{24|\mathcal{S}|} \mathbf{D}_{i,j}, \; \mathbf{D} = |\hat{\mathbf{L}} - \bar{\mathbf{L}}_{\{1,2\}}|.$$

##### 2.2.54 One Minute Scale Heart Rate

***Summary***: Overall one minute scale heart rate estimates. Measured as the number of heart beats per minute [bpm].

***Measure Ids***

1. *bed_heart_rate_1min_scale*

***Type***

1. *Sleep*

**2.2.54.1 Background and Hypothesis** This measure is mostly the same as 2.2.1, however, here estimates are derived by taking the median across 1 minute epochs and the aggregation is based on all 1 minute epochs, instead of per night values. This might capture some distributional aspects that would be lost in 2.2.1.

**2.2.54.2 Description** Here all night records over a pre-defined period are being segmented into one minute epochs. For each epoch, the median heart rate is calculated.

##### 2.2.55 One Minute Scale Respiration Rate

***Summary***: Overall one minute scale respiration rate estimates. Measured as the number of respiration cycles per minute [rpm].

***Measure Ids***

1. *bed_respiration_rate_1min*

***Type***

1. *Sleep*

**2.2.55.1 Background and Hypothesis** This measure is mostly the same as 2.2.5, however, here estimates are derived by taking the median across 1 minute epochs and the aggregation is based on all 1 minute epochs, instead of per night values. This might capture some distributional aspects that would be lost in 2.2.5.

**2.2.55.2 Description** Here all night records over a pre-defined period are being segmented into one minute epochs. For each epoch, the median respiration rate is calculated.

##### 2.2.56 One Minute Scale In-Bed Activity

***Summary***: Overall one minute scale in-bed activity estimates.

***Measure Ids***

1. *bed_activity_1min*

***Type***

1. *Sleep*

**2.2.56.1 Background and Hypothesis** This measure is mostly the same as 2.2.9, however, here estimates are derived by taking the median across 1 minute epochs and the aggregation is based on all 1 minute epochs, instead of per night values. This might capture some distributional aspects that would be lost in 2.2.9.

**2.2.56.2 Description** Here all night records over a pre-defined period are being segmented into one minute epochs. For each epoch, the median in-bed activity is calculated.

#### Acronyms

**AC** arousal count. 32

**DFT** discrete Fourier transform. 15–17, 27

**ERE** eigenbehavior (or PCA) based reconstruction error. 19, 43

**FE** Fridge usage entropy. 18

**IS** intradaily stability. 15
**IV** intradaily variability. 14

**LF** low frequency (0.04–0.15 Hz). 36
**LFHF** power spectral density ratio. 16

**MCI** mild cognitive impairment. 16, 24
**MMSE** Mini Mental State Examination. 15
**MoCA** Montreal Cognitive Assessment. 19

**NBASE** nightly bed activity spectral entropy. 30
**ND** nocturnal dipping. 26
**NHRSE** nightly heart rate spectral entropy. 27, 29
**NRRSE** nightly respiration rate spectral entropy. 29

**PIR** passive infrared. 5–7, 9, 11–22, 24, 30, 39, 43
**PSD** power spectral density. 17, 27

**REM** rapid eye movement. 25, 36, 38, 42, 43

**SE** spectral entropy. 15, 16
**SWT** stationary wavelet transform. 17

**WV** wavelet variance. 16, 17

Thomas Roth. Insomnia: definition, prevalence, etiology, and consequences. *Journal of clinical sleep medicine*, 3(5 suppl):S7–S10, 2007.

John Peever and Patrick M Fuller. The biology of rem sleep. *Current Biology*, 27(22):R1237–R1248, 2017.

Jana R Cooke and Sonia Ancoli-Israel. Sleep and its disorders in older adults. *Psychiatric Clinics*, 29(4):1077–1093, 2006.

Fred Shaffer and James P Ginsberg. An overview of heart rate variability metrics and norms. *Frontiers in public health*, 5:258, 2017.

Altug Didikoglu, Asri Maharani, Gindo Tampubolon, Maria Mercè Canal, Antony Payton, and Neil Pendleton. Longitudinal sleep efficiency in the elderly and its association with health. *Journal of sleep research*, 29(3):e12898, 2020.

Mitsuru Ohishi, Takuro Kubozono, Koji Higuchi, and Yuichi Akasaki. Hypertension, cardiovascular disease, and nocturia: a systematic review of the pathophysiological mechanisms. *Hypertension Research*, 44(7):733–739, 2021.
